# Clinical, neuropathological, and biochemical characterization of ALS in a large CHCHD10 R15L family

**DOI:** 10.1101/2025.09.22.25335938

**Authors:** Justin Y. Kwan, Christian I. Lantz, Vlad A. Korobeynikov, Allison Snyder, Xiaoping Huang, Taryn Haselhuhn, Katherine N. Dore, Angelo Madruga, Laura E. Danielian, Alice B. Schindler, Ruth Chia, Memoona Rasheed, Jody Crook, Marcell Szabo, Makayla Portley, Carolyn M. Sherer, Monique C. King, Tzu-Hsiang Huang, Peter Kosa, Bibiana Bielekova, Michael E. Ward, Chris Grunseich, Neil A. Shneider, Bryan J. Traynor, Derek P. Narendra

## Abstract

Familial forms of ALS are potential candidates for gene-directed therapies, but many recently identified genes remain poorly characterized. Here, we provide a comprehensive clinical, neuropathological, and biochemical description of fALS caused by the heterozygous p.R15L missense mutation in the gene CHCHD10. Using a cross-sectional study design, we evaluate five affected and nine unaffected individuals from a large seven-generation pedigree with at least 68 affected members. The pedigree suggests a high (68 - 81%) but incomplete disease penetrance. Through cloning of the disease-allele from distant members of the family, we establish the disease haplotype in the family. Notably, the haplotype was distinct from that of a previously reported p.R15L mutation carrier with ALS, demonstrating that the variant is in a mutational hotspot. The clinical presentation was notable for being highly stereotyped; all affected individuals presented with the rare ALS variant Flail Arm Syndrome (FAS; also known as, brachial amyotrophic diplegia or Vulpian-Bernhardt Syndrome), suggesting greater involvement of the cervical spinal cord. Consistently, neuropathology from one family member demonstrated substantially increased CHCHD10 protein aggregation and neuronal loss (though absent TDP-43 pathology) in the cervical vs. lumbar spinal cord. This FAS phenotype could be captured by a simple timed finger tapping task, suggesting potential utility for this task as a clinical biomarker. Additionally, through analysis of fibroblast lines from 12 mutation carriers, isogenic iPSC cells, and a knockin mouse model, we determined that CHCHD10 with the R15L variant is stably expressed and retains substantial function both in cultured cells and *in vivo*, in contrast to prior reports. Conversely, we find loss of function (LoF) variants are more common in the population but are not associated with a highly penetrant form of ALS in the UK Biobank (31 in controls; 0 in cases). Together, this argues against LoF and in favor of toxic gain-of-function as the mechanism of disease pathogenesis, similar to the myopathy-causing variants in CHCHD10 (p.G58R and p.S59L). Finally, through proteomic analysis of CSF of variant carriers, we identify that CHCHD10 protein levels are elevated approximately 2-fold in mutation carriers, and that affected and unaffected individuals are differentiated by elevation of two neurofilaments: neurofilament light chain (NfL) and Peripherin (PRPH). Collectively, our findings help set the stage for gene-directed therapy for a devasting form of fALS, by establishing the likely disease mechanism and identifying clinical and fluid biomarkers for target engagement and treatment response.

## Introduction

Amyotrophic Lateral Sclerosis (ALS) is a relentlessly progressive disorder, resulting from degeneration of motor neurons in the motor cortex and spinal cord ^1,2^. Gene directed therapies are promising for familial forms of ALS (fALS) caused by mutations in single genes ^3–5^. For these therapies to be effective, however, the disease mechanism must be known, and biomarkers must be identified to track target engagement and treatment response. While these are well-established for some genetic causes of fALS, they remain under-explored for many recently identified fALS genes, including *CHCHD10* ^6^.

Mutations in *CHCHD10* were first identified as a dominant cause of fALS in 2014 ^7^, with segregation shown for two variants, p.S59L and p.R15L, in multiple families ^7–10^. The p.R15L causes a pure ALS phenotype, whereas the p.S59L variant is clinically variable with ataxia, frontotemporal dementia, and adult-onset mitochondrial myopathy also reported in some patients. Other variants in *CHCHD10* cause distinct neuromuscular disorders including autosomal dominant isolated mitochondrial myopathy (IMMD) (p.G58R) and a late-onset form of spinal muscular atrophy named SMA-J for Jokela type (p.G66V) ^11–13^. A dose effect has been observed for two of these variants (p.S59L and p.G66V) *in vivo*, with a more severe phenotype observed for homozygotes compared to heterozygotes ^14,15^. Only one neuropathological case of CHCHD10 has been reported for a patient with the p.R15L variant ^16^. The case was notable for aggregates of CHCHD10 in the spinal cord and brain and lack of TDP-43 pathology.

*CHCHD10* encodes a small mitochondrial protein. Together with its paralog CHCHD2, CHCHD10 supports the inner mitochondrial membrane (IMM) structure to maintain oxidative phosphorylation (OXPHOS) (reviewed in ^17^). Surprisingly, loss of one or both paralogs is well-tolerated in mice, suggesting that they may be dispensable for mammalian life ^18–20^. While a toxic gain-of-function (GoF) mechanism is established for some *CHCHD10* mutations causing myopathy ^13,14,21^, the disease mechanism remains less certain for motor neuron impairment caused by CHCHD10 mutations, with studies supporting both loss of function (LoF) ^14,22–24^ and GoF models ^14,21,25,26^ for motor neuron loss. The pathogenicity of the *CHCHD10* p.R15L variant has also been challenged, on account of the small families initially reported and the presence of unaffected mutation carriers in the pedigrees ^27^.

Here, we provide a comprehensive assessment of the p.R15L variant in a 7-generation US family in which 68 members were affected with ALS. Using a combination of clinical, neuropathological, genetic, and cell and mouse modeling approaches, we make several key findings. First, we establish that the *CHCHD10* p.R15L mutation causes an uncommon ALS variant called Flail Arm Syndrome (FAS) (also known as, brachial amyotrophic diplegia or Vulpian-Bernhardt Syndrome). Through neuropathological evaluation of one family member, we demonstrate that this pattern correlates with the burden of CHCDH10 protein aggregation within the spinal cord, with greater pathology observed in the cervical vs. lumbar spinal cord. This distinguishes the p.R15L phenotype from the other well-characterized motor neuron disease-causing variants in *CHCHD10*, p.S59L and p.G66V, and other forms of fALS. Second, we demonstrate that the endogenous gene product of the *CHCHD10* p.R15L variant retains substantial protein stability and function in iPSC and mouse models. This demonstrates that the variant is a hypomorph and not LoF, as proposed previously. We additionally establish that heterozygous LoF variants in *CHCHD10* are not a highly penetrant cause of ALS in the UK Biobank (UKBB), which further argues against a LoF mechanism of pathogenesis. Fourth, through proteomic analysis of the CSF, we identify that the CHCHD10 protein is elevated by approximately two-fold in both symptomatic and presymptomatic mutation carriers relative to other forms of ALS and healthy controls. This suggests mutant CHCHD10 is cleared from CSF less efficiently than WT CHCHD10 *in vivo* and is consistent with a toxic GoF model. Finally, we establish that neurofilament light chain (NfL) and Peripherin (PRPH) readily distinguish presymptomatic from symptomatic mutation carriers and may thus serve as biomarkers for phenoconversion and therapeutic response. Together, these findings demonstrate that the *CHCHD10* p.R15L variant causes a distinct form of fALS, most likely by a GoF mechanism, setting the stage of gene-directed therapy.

## Materials and Methods

### Characterization of affected and unaffected individuals with CHCHD10 variants

Study participants were recruited and evaluated in the Neurogenetics and/or Neurodegenerative Disorders Clinic at the National Institute of Neurological Disorders and Stroke (NINDS) Intramural Research Program in 2022 - 2024. One participant was seen remotely. Genetic status was confirmed in all participants. All participants with the CHCHD10 p.R15L mutation were eligible to participate in the study. All participants gave written informed consent according to the Declaration of Helsinki to protocols approved by the Institutional Review Board of NIH before undergoing research procedures (NCT03225144 and NCT00004568). A board-certified neurologist with specialized training in neuromuscular disorders performed comprehensive neurological evaluations in all patients. Quantitative measures of motor function (articulatory coordination [verbal diadochokinesis or PaTaKa testing], finger and foot tapping speed, instrumented timed up and go [iTUG] test, and 25-feet gait evaluation) and cognitive evaluation using the ALS Cognitive Behavioral Screen (ALS-CBS) and a standard battery of neuropsychological tests were performed in all participants ^28^. The iTUG test and 25-feet gait evaluation were assessed using the APDM Mobility Lab® Opal V2R inertial sensor system worn on the feet, wrists, and trunk (Version 5, Portland, OR). The revised ALS Functional Rating Scale (ALSFRS-R) and CDR® NACC-FTLD-M were used as measures disease severity ^29–31^. A 3T magnetic resonance imaging (MRI) scanner (Philips Achieva, Best, the Netherlands) with a receive-only, eight-channel Sense head coil was used for imaging in all participants. A high-resolution T1-weighted sequence (magnetization-prepared rapid gradient echo, TE = 3.77 ms, TR = 8.17 ms, FOV 240 x 240, Resolution 256 x 256, 181 slices, 1 mm thickness, voxel = 0.9375 x 0.9375 x 1 mm) was acquired for volumetrics and cortical thickness measurements. Imaging processing was performed with FreeSurfer imaging analysis suite (http://surfer.nmr.mgh.harvard.edu/) using previously described and validated procedures ^32–34^. Images were visually inspected for errors in segmentation, which were manually corrected. The tessellated surfaces were inflated and registered to the Desikan-Killiany atlas which parcellates the cerebral cortex into 34 regions in each hemisphere ^35^. The total intracranial volume (TIV) was calculated using Statistical Parametric Mapping 12 ^36^. Cortical gray matter volumes are expressed as a percentage of the TIV to account for differences in the head size. Blood, CSF, and skin were obtained from participants for clinical evaluation and/or research seen under the protocol unless refused by the participant or a contraindication existed. SomaScan® and Simoa® assays were performed in house. The Olink® Explore HT assay was performed commercially by Psomagen. For analysis of *CHCHD10* variants in the UKBB, data was accessed through the UK Biobank Research Analysis Platform (https://www.dnanexus.com/partnerships/UKBiobank). ALS cases were defined as individuals diagnosed with ICD-10 code G12.2. To define a neurologically healthy control group, we excluded samples with ICD-10 codes corresponding to other neurodegenerative and muscle disorders: AD (any codes G30-G32), PD (G20-G26), dementia-related (F00-F05), disease of myoneural junction and muscle (G70-G73), or polyneuropathies and other disorders for the peripheral system (G60-G64). A variant was considered LoF if it was annotated as predicted LoF (pLoF) for the MANE Select CHCHD10 transcript (ENST00000484558.3 / NM_213720.3) in gnomAD v4.1 ^37^.

For the neuropathological examination of participant VI-10, five-micron tissue sections were deparaffinized in xylene and rehydrated in ethanol solutions followed by antigen retrieval by autoclaving the slides in Tris-EDTA buffer (10 mM Tris pH 9, 1 mM EDTA, 0.05% Tween 20) for 10 min. The slides were incubated with the CHCHD10 primary antibody (Sigma Aldrich, clone 11F11.2, MABN1524,1:300) in TBS solution with 0.01% Triton X-100 and 5% normal goat serum overnight at 4 °C. The slides were subsequently incubated with HRP-conjugated secondary antibodies (SuperBoost™ Goat anti-Mouse Poly HRP, Invitrogen, B40961),developed with DAB Substrate Kit (Thermo Fisher Scientific) for 10 min, and counterstained with Mayer’s hematoxylin for 2 min."

### Establishment and culture of primary dermal fibroblasts lines

Patient fibroblast lines were established from 3-mm punch skin biopsies taken from the forearm. Fibroblasts were cultured in high glucose DMEM, pyruvate, L-glutamine, 10% FBS with pen/strep. Cell lines were assayed at passage 11 or before. The transformed human fibroblast line from Canadian Individual 1, carrying a heterozygous p.R15L variant, was a kind gift from Eric Shoubridge. The cell line and the clinical course for this participant have been described previously ^16,23^.

### Establishment of CHCHD10 variant iPSC lines

The *CHCDH10* R15/R15L, KO, and compound heterozygous indel (c.270-299del; c.288delA) line was created by nucleofection with HiFi Cas9 ribonucleoprotein (RNP). Briefly, WTC11 iPSCs (gift of Dr. Bruce R. Conklin (Gladstone Institute of Cardiovascular Disease, UCSF)) were electroporated with HiFi Cas9 nuclease V3 (IDT, 1081061), a small guide RNA (sgRNA) targeting *CHCHD10* (sequence in supplementary table), with or without a single-stranded template oligonucleotide (ssODN) for homology-directed repair (HDR) (sequence in supplementary table). Reagents and cell mixtures were prepared using the P3 Primary Cell 4D X Kit L (Lonza, V4XP-3024) and nucleofection was accomplished with the Lonza 4D-Nucleofector Core Unit (Lonza, AAF-1003B) nucleofector. Isolated single cells were grown to produce a monoclonal line. Sanger sequencing confirmed HDR or insertion/deletion in selected clones. The differentiation cassette was integrated into the genome of the isogenic series of iPSC cell lines described using a PiggyBac system. The N50-PB-tri-TO-NIL-BFP-puro plasmid was co-transfected with the K13-EF1a transposase with Lipofectamine Stem Transfection Reagent (Invitrogen, STEM00003). Transfected cells were selected with a range of 0.5-2 μg/mL puromycin (Invivogen, ant-pr-1).

### iPSC Culture and Differentiation

iPSCs were cultured in Essential 8 medium (E8) (Gibco, A1517001) on plates coated with Matrigel (Corning, 354277). After passaging or thawing cells, media was supplemented with 10 μM Y-27632 ROCK inhibitor (Tocris Bioscience, 1254) for 24 hours to improve cell survival. Before differentiating to iLMNs, cells were dissociated to single-cells with Accutase (Gibco, A1110501) and plated onto a Matrigel-coated 10 cm plate in E8 medium supplemented with ROCK inhibitor. Starting on day 0, cell media was replaced with Neural Induction Medium (NIM) consisting of DMEM/F12 (Gibco, 11320033), B27 supplement (Gibco, 17504044), GlutaMAX supplement (Gibco, 35050061), non-essential amino acids (Gibco, 11140050), ROCK inhibitor, compound E (Sigma-Aldrich, 565790), and doxycycline (Sigma-Aldrich, D9891). On day 2, cells were dissociated with Accutase and transferred to plates coated with 0.1% poly(ethyleneimine) (Sigma, P3143). Cells were plated in Neural Differentiation Medium (NDM) consisting of DMEM/F12, BrainPhys (StemCell Technologies, 05790), B27 supplement, GlutaMAX supplement, non-essential amino acids, ROCK inhibitor, compound E, Laminin (Gibco, 23017015), BDNF (PeproTech, 450-02-10UG), GDNF (PeproTech, AF-450-10-10UG), NT-3 (PeproTech, 450-03-10UG), Culture One supplement (Gibco, A3320201), and doxycycline. On day 2, cells were plated in NDM supplemented with 40uM 5-Bromo-2′-deoxyuridine (BrdU; B9285, Sigma-Aldrich) for 24 hours, then on day 3, cell media was replaced with NDM without BrdU supplement. On day 4, cell media was replaced with Neuron Medium (NM) consisting of NDM media without DMEM/F12 and non-essential amino acids. On day 7, half-media changes of NM were done and continued every 3 days until differentiation day 9.

### Affinity-Purification Mass Spectrometry (AP-MS) of CHCHD10

*CHCHD10* wildtype (WT), indel, and R15L iPSCs were collected from 10cm plates in 4 technical replicates per line. Each replicate was solubilized in an affinity purification (AP) lysis buffer containing 20 mM Bis-Tris pH 7, 50 mM NaCl, 10% (v/v) glycerol and 1% digitonin, plus Protease Inhibitor Cocktail (Sigma; P 8340). Lysates were incubated on ice for 30 mins and then centrifuged at 21300 g for 10 mins. Supernatant samples were kept on ice. Immunoprecipitations were performed according to manufacturer’s protocol. Briefly, the Pierce™ Protein A Magnetic Beads (Thermo Scientific; 88846) were prepared by washing two times with AP buffer and resuspending in AP buffer with 1 mM EDTA. The supernatant samples were applied to the prepared beads and CHCHD2/CHCHD10 custom antibody (Genscript; SC1809) and incubated for 1.5 hr at room temp on a rotator. Bound proteins were washed with AP buffer containing 0.001% digitonin two times and MQ water one time for 5 mins on ice and eluted using Laemmeli buffer containing 1.05% SDS, 13% glycerol (w/v), 5% 2-mercaptoethanol, 0.005% bromophenol blue and 32.9 mM Tris HCl, pH 6.8. The eluted proteins were heated at 95 for 5 mins and kept at -80 before submitting for proteomics.

### Confocal Microscopy of CHCHD10 variant iLMN lines

Induced lower motor neuron-like cells (iLMNs) and iPSCs were fixed with 4% paraformaldehyde in PBS, permeabilized with 0.25% Triton-X in PBS, blocked with 1% BSA in PBS, and stained with primary antibodies at concentrations indicated below in 1% BSA in PBS overnight, then incubated with secondary antibodies at a concentration of 1:1000 in 1% BSA in PBS for one hour.

The following primary antibodies were used: cytochrome c (BD Biosciences, 556432) (1:500), CHCHD10 (Sigma, HPA003440) (1:500), TUBB3 (BioLegend, 801201) (1:500), and HB9 (DSHB, 81.5C10) (1:50). Where indicated, cells were co-stained with DAPI (Thermo Scientific, 62248) (1:5000 in PBS).

### Sequencing CHCHD10 R15L haplotypes

Genomic DNA was extracted from fibroblast lines of the individuals indicated. Long range PCR was used to amplify the *CHCHD10* gene. The gene was TOPO cloned into the pCR4-TOPO TA vector using the TOPO TA Cloning Kit for sequencing (Invitrogen, 450030) following the manufacturer’s instructions. Single bacterial colonies were selected after ligation and grown up. Each clone contains only one of the two alleles amplified by long range PCR. DNA was isolated from each using the QIAprep Spin Miniprep Kit (Qiagen, cat#27104). Several sequencing primers were used to generate overlapping Sanger sequencing reads of each clone. A consensus sequence for the haplotype of each allele was determined by manual inspection of all clones obtained from each sample.

### Establishment of Chchd10^R15L^ knock-in mouse line

Mouse work was performed under the NINDS ACUC protocol ASP1395. The *Chchd10^hR15L/+^* KI mouse line was generated using CRISPR-Cas9 aided homologous recombination. A donor vector was commercially synthesized (Genescript) containing the coding region of the human *CHCHD10* p.R15L allele determined by cloning and Sanger sequencing above as well as approximately 1 kb flanking regions corresponding to the mouse Chchd10 locus (sequence in supplemental material). Gene replacement was carried out in a F1 hybrid of C57Bl6 and SJL by electroporation of the donor vector together with a gRNA targeting the mouse *Chchd10* locus. The sequence of donor vector is in the supplemental document file. KI of the transgene was confirmed by Sanger sequencing at each junction site and at the mutation site. To determine whether the allele can replace *Chchd2* and *Chchd10* function, the mice were crossed with *Chchd2^-/-^*; *Chchd10^-/-^*mice described previously ^20^. The *Chchd10^G58R/+^*and *Chchd10^S59L/+^* KI mouse models were described previously ^13,20^. All animals tested were included in the results; none were excluded. No exclusion criteria were set. No randomization was performed as there was no intervention or assays sensitive to experimenter intervention such as behavioral analysis. Similarly, no blinding was used. The primary outcomes tested were the pattern of CHCHD10 expression and its localization, and activation of the mitochondrial ISR by measuring the following protein markers by immunoblotting (OPA1, OMA1, MTHFD2, PYCR1, PSAT1, P5CS). Additionally, body weights were tracked.

### Fractionation of cell and tissue lysates and immunoblotting

For immunoblotting of cells, cell pellets were lysed in RIPA buffer on ice for 30 min and then cleared by centrifugation at 21,130g. Protein concentration was determined using the BCA assay. Mitochondria were isolated from the mouse hearts as described previously^13^. For soluble/insoluble fractionation, 30 μg of mitochondrial and cytosolic fractions were treated with 1% TX-100 in 50 mM Tris, and after 21,300 g centrifugation for 10 minutes the supernatant was used as the soluble fraction and the pellet was treated again with 2% SDS in 50 mM Tris buffer at room temperature for about 25 minutes. The supernatant was used as the insoluble fraction. All material from the soluble and insoluble fractions was loaded onto AnyKD Criterion TGX precast midi protein gels (Biorad) and proteins were transferred to nitrocellulose membranes. Membranes were blotted. Densitometry measurements were performed using Fiji (NIH) and Image Studio (LICOR). OMA1-cleaved S-OPA1 bands were calculated by measuring the maximum intensity of each of the five bands in a linescan of the optical density of the five OPA1 bands on the blot using Fiji. After subtracting background intensity, the peaks of the c and e bands were summed and divided by the sum of the five bands (a–e) to obtain the percentage of OMA1-generated S-OPA1 from total OPA1. The following antibodies were used for immunoblotting: OPA1 (BD science #612606), CHCHD10 (Sigma #HPA003440), CHCHD2 (Proteintech #66302-1-Ig), GAPDH (Proteintech #60004-1-Ig), total OXPHOS (Abcam #ab110411), OMA1 (Santa Cruz #SC-515788), MTHFD2 (Proteintech #12270-1-AP), HSP90 (Proteintech #13171-1-AP), P5CS (Proteintech #177191-1-AP), PSAT1 (Proteintech #10501-1-AP), PYCR1 (Proteintech, #13108-1-AP).

### Clinical proteomics

Cerebrospinal fluid proteomic data on the Olink Explore HT platform (Thermo Fisher Scientific Inc., Waltham, MA) was generated by Psomagen, a Contract Research Services vendor. Samples used were from healthy controls, sporadic ALS cases (negative gene testing), ALS cases due to a *C9orf72* expansion, ALS cases due to the *CHCHCD10* p.R15L variant, or presymptomatic *CHCHD10* p.R15L variant carriers. In the Olink assay, capture and detection antibodies for each analyte are conjugated to oligonucleotides with barcodes and pooled together, allowing highly multiplexed measurement of proteins in the sample. Capture and detection of an analyte by the antibody pairs will allow for DNA amplification that can be measured by next-generation sequencing. All samples were run on the same plate and their order was randomized. Recommended initial processing steps were followed in the Olink software to exclude analytes not passing the quality control checks. We used the PC Normalized Protein eXpression (NPX) transformed values in our analysis. These represent the log_2_ transformed ratio of sample assay counts to extension control counts.

Cerebrospinal fluid proteomic measurements on the SomaScan 7K platform were generated from ALS cases with no variant or family history (sporadic ALS), a *C9orf72* expansion, or the *CHCHCD10* p.R15L variant, presymptomatic individuals with the CHCHD10 p.R15L variant, or healthy controls. SomaScan is a single-stranded DNA aptamer-based proteomics platform. Some of the SomaScan data were used in a prior publication ^38^. The cases matched those used in the Olink analysis, except that one sALS patient sample was substituted due to sample availability. These same cases samples were also analyzed by Simoa for neurofilament light chain (NfL). Controls were matched 1:1 to *CHCHD10* p.R15L carriers as follows: each variant carrier was matched to a healthy donor of the same sex and closest age within ±5 years; donors were used once (no replacement); we matched “hardest first” (people with fewest eligible donors) to minimize dropouts. Matching used a custom function implementing exact matching on sex and nearest-age selection under a ±5-year caliper with no replacement and scarce-first ordering; reproducibility was ensured with seed = 123. The healthy volunteer samples were different from those used in the Olink analysis. The RFU values were log_2_ transformed and used in downstream analysis without further adjustment.

### Immunofluorescence of mouse tissues

For analysis of tissue, mice were anesthetized with isoflurane and transcardially perfused with 25 mL PBS (for heart tissue) or 4% PFA (for brain and spinal cord). For heart tissue, the heart midventricular region was dissected and immunostained for CHCHD10 and cytochrome *c* as described previously ^13^. For brain and spinal cord tissue, Compresstome sections of the tissue were immunostained as follows: tissues were sectioned at 35 µm using the Compresstome VF-510-0Z (Precisionary). Free floating sections were obtained and washed with PBS twice for 5 minutes each and in 0.1%Triton X-100 in PBS (PBST) twice for 10 minutes each. Sections were then blocked in 0.4% PBST with 10% Normal donkey serum (NDS) for 2 hours at room temperature. Tissues were placed with primary antibodies overnight in 0.3% PBST with 3% NDS. The next day, tissues were washed with 0.1% PBST 4 times for 10 minutes each and then incubated with secondaries in 0.1% PBST for 1-2 hours at room temperature. Tissues were washed with 0.1% PBST twice for 10 minutes each and with PBS 2x for 5 minutes each at room temperature. Counter staining was done with either DAPI or blue, fluorescent Nissl for 10 minutes. All tissues were mounted on slides and coverslipped with prolong diamond mounting media and sealed with nail polish. Images were obtained using Olympus FLUOVIEW FV3000 confocal laser scanning microscope.

### Statistical Analysis

Visualization and statistical analysis for data in dot plots and bar graphs was performed in Graphpad Prism 10. The specific tests performed are indicated in the Figure Legends. For the clinical proteomics datasets, statistical analysis and visualization were performed using custom Python scripts. For each comparison, an ANOVA was performed using the Scipy (1.16.1) library followed by a Benjamini Hocherg false discovery rate (FDR) correction for multiple comparisons using the statsmodels (0.14.4) library. For pair-wise comparisons, a two-sided Student’s t-test was used followed by an FDR correction for multiple comparisons. A protein was determined to be significant in the pairwise comparison if it was significant both across groups using the corrected ANOVA p-value and significant for the specific pairwise comparison using the corrected t-test value. Annotation of mitochondrial proteins was performed using a custom python script with annotations drawn from the MitoCarta3.0 dataset ^39^. CHCH-domain containing proteins were annotated using the following list: [’CHCHD1’, ’MIC19’, ’CHCHD3’, ’CHCHD6’, ’MIC25’, ’CHCHD4’, ’MIA40’, ’CHCHD10’,’NDUFA8’, ’NDUFB7’, ’NDUFS8’, ’UQCRH’, ’QCR6’, ’COX6B1’,’COX6B2’,’COX17’, ’COX19’, ’CHCHD7’, ’COX23’, ’COA4’, ’COA5’, ’PET191’, ’COA6’, ’CMC1’, ’CMC2’,’TRIAP1’,’MDM35’,’CHCHD5’,’CMC4’,’UPF0545’,’C17orf89’]. For analysis of AP-MS proteomics data, data was first filtered for mitochondrial proteins using human MitoCarta3.0. Statistics were performed using Perseus 1.6.14.0: missing data was imputed with “Replace missing values from normal distribution" function; a two-sided Student’s t-test was then performed using default settings with absolute log_2_ fold change over 1; finally, a permutation-based FDR was used for multiple comparisons. Data was visualized using a custom Python script with the Matplotlib and Seaborn libraries. No sensitivity analysis was performed.

### Data Availability

Mass spectrometry based proteomics data will be deposited in the MassIVE public repository. Source data for confocal images, immunoblotting images, and numerical data used to generate graphs will be deposited in BioStudies. Source MRI images included identifiable information and so were not included. Raw values for Olink and Somascan clinical proteomics (NPX and RFU values, respectively) are included in Supplemental Tables S4 and S5.

## Results

### Genetic analysis confirms a shared CHCHD10 haplotype in large ALS pedigree around the p.R15L mutational hotspot

At the NIH Clinical Center, we evaluated five affected and nine unaffected *CHCHD10* p.R15L heterozygous mutation carriers from three extended families (including one previously reported as USALS#5) ^9^. Genealogical records showed that the three extended families were distantly related though a common ancestor, who immigrated to the US from Germany early in the 19^th^ century. In the extended pedigree, eight affected members were known carriers of the heterozygous *CHCHD10* p.R15L variant, while no affected family member tested negative (Fig. 1A and S1A).

**Figure 1.**
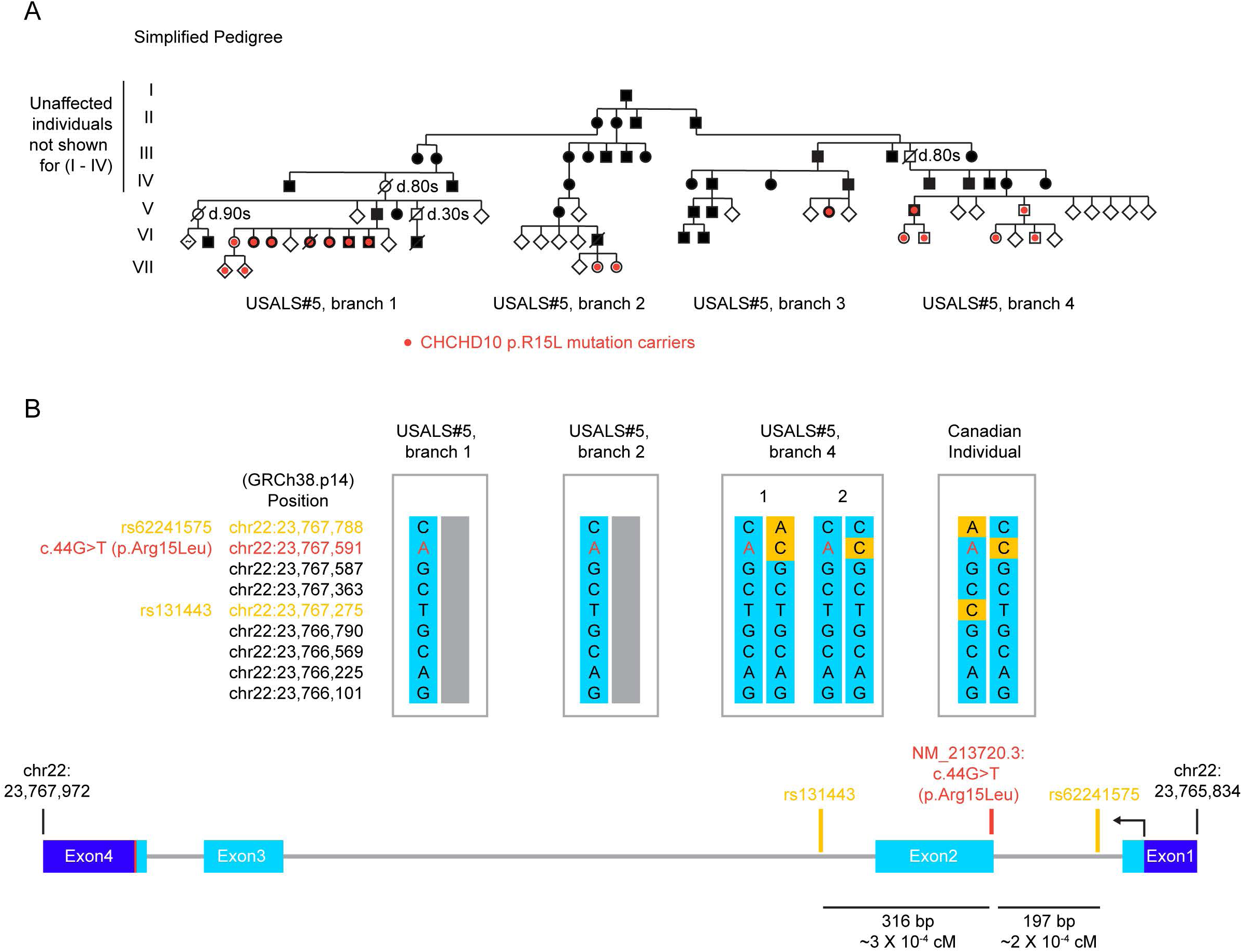
Pathogenic CHCHD10 R15L variant in large family with familial ALS. (A) Simplified pedigree shows segregation of the heterozygous CHCHD10 p.R15L variant in a large family. Solid black symbols indicate those affected with ALS. Red dots indicate those testing positive for the heterozygous CHCHD10 p.R15L variant. (B, top) Common polymorphisms surrounding the pathogenic CHCHD10 p.R15L variant (red font) are shown for the indicated individuals from branches of the USALS#5 family and a sporadic ALS patient that was previously reported (Canadian Individual 1) ^16^. Haplotype was established by topo cloning of each allele following long-range PCR of genomic *CHCHD10* and Sanger sequencing. Common single nucleotide polymorphisms (SNPs) not matching the haplotype of the USALS#5 have an orange background. The haplotype surrounding p.R15L for Canadian Individual 1 is distinct from that surrounding distantly related members of the USALS#5 family. (B, bottom) the location of two common SNPs surrounding p.R15L are indicated (orange).

Cloning and sequencing of the *CHCHD10* disease allele from members of three of the branches confirmed that the rare p.R15L variant is part of a shared haplotype (Fig. 1B and S1B). Individuals carrying this haplotype were separated by as many as five generations and so were estimated to share less than 1% of variable autosomal DNA, strongly supporting that p.R15L is the pathogenic variant. This shared haplotype was not present in an unrelated Canadian patient with sporadic ALS and the p.R15L variant ^16,23^ (Fig. 1B and S1B, Canadian Individual 1). This marks the p.R15L position as a mutational hotspot, with the mutation arising more than once on different haplotypes.

Altogether, the extended pedigree included at least 68 individuals with ALS and 32 sibling groups with at least 1 affected sibling (Fig. S1A). Disease penetrance in the family was high but incomplete. 81% (26/32) of affected sibling groups had an affected parent (Table S1). Similarly, 34% (68/199) of individuals in each affected sibling group had ALS. This is 68% of what would be predicted by a full penetrance model (i.e., where 50% of siblings are affected) (Table S1). Among the obligate carriers were three who died after 80 years without motor symptoms, per first-hand accounts of family members interviewed. Together, this suggests a high penetrance in the range of 68 – 81%.

Among those in the extended pedigree, a first-hand account of the clinical course was available for 17 (Table S1). The median onset of ALS in this group was 58 years (range 35 – 71, N=17) with a median disease duration of 8 years (range 4 – 18, N=11). The pattern of motor symptom onset and progression was highly stereotyped in each. Weakness occurred first in an upper extremity, followed by weakness in the contralateral upper extremity, before progressing to inspiratory weakness and/or bulbar symptoms. Weakness in the lower extremities was a late occurrence or absent in each case. Dementia was not diagnosed for any of the participants with ALS with one exception per the recollections of family members. In contrast to others in the family, the initial symptoms for this individual were cognitive, and he was clinically diagnosed with Parkinson’s disease and Alzheimer’s disease in addition to ALS during his disease course. This raises the possibility of a comorbid common neurodegenerative disorder in this individual.

### *CHCHD10* p.R15L causes flair arm syndrome, an ALS subtype, with incomplete penetrance

The flail arm pattern of weakness was confirmed by clinical evaluations at the NIH in five affected family members, who were descendant from three distant family branches. The participants were evaluated two to five years after their first motor symptom onset. Their ALSFRS-R scores ranged from 30 to 38, indicating mild to moderate disease severity. In all participants, arm muscles were the weakest on confrontational motor testing with a proximal to distal gradient (Fig. 2A). Except for mild iliopsoas weakness in one individual, all participants retained 5/5 strength (on the Medical Research Council (MRC) scale) in their major lower extremity muscles groups. Electrophysiological testing in all symptomatic gene mutation carriers showed findings consistent with both active and chronic denervation, most pronounced in the cervical myotomes, meeting the electrophysiologic criteria for diffuse motor neuron disease. Signs of a primary myopathic process were absent. In contrast to the affected participants, clinical examinations were normal for the nine unaffected participants, including an unaffected participant in her 70s who had affected younger siblings, pointing to a discrete disease onset. Together, these findings establish that each symptomatic participant has FAS.

**Figure 2.**
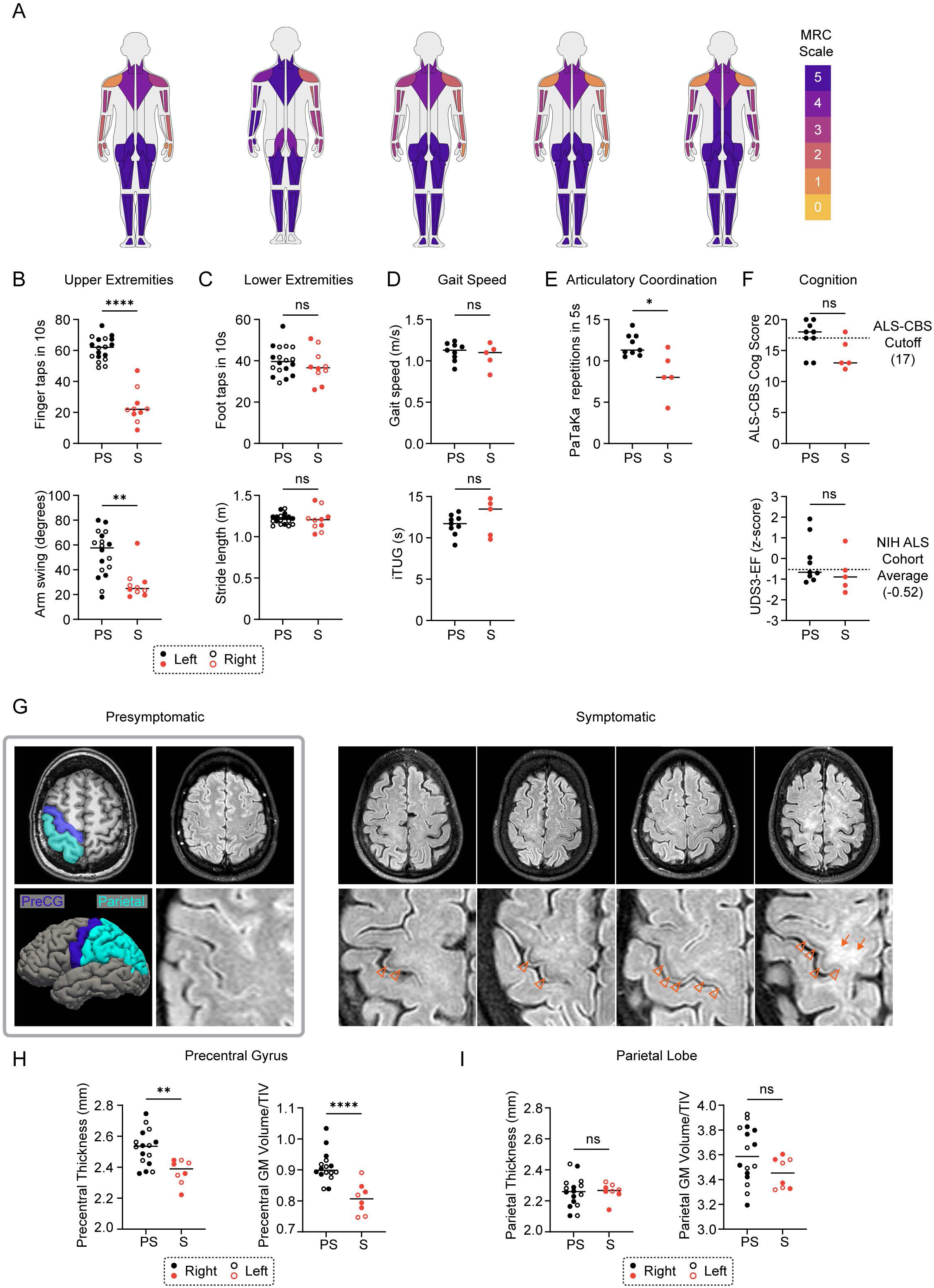
CHCHD10 R15L ALS presents clinically as Flail Arm Syndrome. (A) Pattern of weakness on confrontational motor testing in affected participants is represented by MuscleViz visualization ^58^. Scoring follows the Medical Research Council (MRC) scale for muscle strength grading. (B) Upper extremity motor function for presymptomatic (PS) and symptomatic (S) *CHCHD10* p.R15L mutation carriers, as measured by the number of finger taps in 10 seconds (top) and degree of arm swing during 25-foot instrumented walk over 4 trials (bottom). (C) Lower extremity motor function as measured by the number of foot taps in 10 seconds (top) and length of stride during 25-foot instrumented walk over 4 trials (bottom). In (B and C) data from left and right extremities appear as solid and open data points, respectively. (D) Gait speed on a 25-foot instrumented walk over 4 trials (top) and instrumented timed up and go (iTUG) (bottom). Gait speed is the average of 4 trials and iTUG is the average of two trials. (E) Articulatory coordination as measured using the PaTaKa speech test. Number of repetitions of the syllables Pa, Ta, and Ka in five seconds is represented. Two trials were averaged. (F) Cognition (top) and executive function (bottom) measured using the ALS-CBS and UDS3-EF, respectively. Dotted line (top) indicates the cutoff value for normal (≥ 17) in this assessment ^59^. Dotted line (bottom) indicated the average score in the NIH ALS cohort. (G) Axial T1-weighted MRI and FreeSurfer brain reconstruction highlighting the precentral gyrus (dark blue) and parietal lobe (light blue). Axial T2-weighted MRI from a presymptomatic participant (gray box) and four symptomatic participants. Precentral gyrus hypointensity (arrowhead) and adjacent white matter hyperintensity (arrow) is observed. (H) Precentral gyrus thickness (left) and gray-matter volume as a percentage of the total intracranial volume (TIV) (right). Data from right and left hemispheres appear as solid and open datapoints, respectively. (I) Parietal cortex gyral thickness (left) and parietal gray matter volume as a percentage of the TIV (right). Data from right and left hemispheres appear as solid and open datapoints, respectively. In all panels, ns, *, **, ***, ***” correspond to not-significant, p ≤ 0.05, p ≤ 0.01, p ≤ 0.001, and p ≤ 0.0001, respectively. Statistics were calculated using the Mann-Whitney test.

Quantitative motor testing further captured the selective involvement of the upper extremity muscles observed in FAS. Arm swing angle was significantly reduced on instrumented gait measurements (Fig. 2B and Table 1), whereas gait speed, iTUG, and stride length were similar to presymptomatic participants (Fig. 2C and D and Table 1). Finger tapping speed, but not foot tapping speed, was similarly reduced (Fig. 2B and C). Of these quantitative motor measures, finger tapping speed was the clinical measure that distinguished symptomatic participants and may be useful for tracking disease progression and treatment response, particularly early in the disease course. By contrast, gait-related measures like iTUG were insensitive in this population. Articulatory coordination was also reduced in many symptomatic participants, consistent with bulbar dysfunction (Fig. 2E and Table 1). Approximately half of FAS patients develop bulbar symptoms; consistent with this, articulatory coordination was markedly slowed in one participant who had more severe bulbar dysfunction ^40^. Dysarthria and dysphagia were reported by two of the five affected individuals, but both had normal evaluations on video fluoroscopic swallowing examination.

**Table 1.** CHCHD10 Demographics and Clinical Features.

|  |  | Presymptomatic | ALS |
| --- | --- | --- | --- |
| Male:Female |  | 4:5 | 2:3 |
| Age, years |  | 40.89 (14.10) | 64.00 (3.08) |
| Symptom Duration, months |  | - | 60.80 (15.69) |
| ALSFRS-R |  | 48.00 (0.00) | 35.00 (3.00) |
| Finger tapping, taps/10 secs | Right | 64.34 (6.49) | 24.14 (14.21) |
|  | Left | 58.03 (7.47) | 23.40 (8.22) |
| Foot tapping, taps/10 secs | Right | 40.51 (5.97) | 39.58 (6.04) |
|  | Left | 39.44 (8.28) | 35.46 (8.89) |
| Gait speed, m/sec |  | 1.11 (0.11) | 1.06 (0.15) |
| Instrumented Timed Up and Go, secs |  | 11.59 (1.25) | 12.50 (2.27) |
| Articulatory Coordination, repetitions/5 secs |  | 11.91 (1.31) | 8.39 (2.75) |
| MMSE |  | 28.11 (2.32) | 27.60 (0.89) |
| ALS-CBS, Cognitive |  | 17.22 (2.64) | 14.40 (2.51) |
| ALS-CBS, Behavioral |  | 44.78 (0.67) | 41.60 (4.77) |
| Frontal Behavioral Inventory |  | 0.11 (0.33) | 2.00 (2.92) |
| Letter Fluency – “F”* |  | 15.67 (4.64) | 11.00 (3.32) |
| Categorical Fluency – “Animals”* |  | 24.44 (6.29) | 19.60 (4.28) |
| HVL T Delayed Recall* |  | 11.00 (1.32) | 10.20 (1.30) |
| BVM T Delayed Recall* |  | 9.56 (3.21) | 10.25 (4.66) |
| UDS3-EF |  | -0.14 (0.96) | -0.71 (1.10) |
| CDR® plus NACC FTLD, SB |  | 0 | 0.10 (0.22) |
| CDR® plus NACC FTLD, global score |  | 0 | 0.10 (0) |
| CDR® NACC FTLD-M, SB |  | 0 | 2.10 (0.23) |
| CDR® NACC FTLD-M, global score |  | 0 | 1.00 (0) |

Values listed as mean (SD)
\*Raw scores reported
ALSFRS-R = ALS Functional Rating Scale, revised; MMSE = Mini Mental Status Examination; ALS-CBS = ALS Cognitive Behavioral Screen; HVL T = Hopkins Verbal Learning Test; BVM T = Brief Visuospatial Memory Test; UDS3-EF = Uniform Data Set v3.0 EF composite score; CDR® plus NACC FTLD = CDR Dementia Staging Instrument PLUS National Alzheimer’s Coordinating Center Frontotemporal Lobar Degeneration; SB = Sum of Boxes

In symptomatic participants, the disease severity, as determined by the CDR® NACC FTLD-M, was mild and driven by motor impairment (Table 1). None of the symptomatic participants met clinical criteria for mild cognitive impairment or dementia. There was no significant difference in performance on visual or verbal memory tasks (Table 1). However, there was qualitatively lower performance on measures of executive function, including phonemic fluency, and the Uniform Data Set v3.0 executive function compositive score (UDS3-EF) (Fig. 2F and Table 1). Additionally, two presymptomatic and four symptomatic participants scored in the impaired range on the ALS-CBS (Table 1).

In symptomatic participants, clinical MRI of the brain showed precentral gyrus hypointensity and adjacent white matter hyperintensity on T2-weighted sequence due to cortical motor neuron degeneration (Fig. 2G) ^41,42^. Quantitative MRI measures also revealed changes consistent with upper motor neuron (UMN) involvement. Compared to the presymptomatic participants, the symptomatic participants had thinning of the precentral gyrus (PCG) cortex (Fig. 2G and H). There was no difference in the adjacent parietal lobe cortical thickness and no substantial atrophy was present in other frontotemporal cortical regions, suggesting that it was focal to the PCG (Fig. 2I).

Taken together, the clinical assessment suggests a stereotyped pathological process that, in addition to cortical motor neuron degeneration, starts in the cervical spinal cord, progresses to the lower brain stem, and ultimately reaches the lumbar spinal cord in the later stages of the disease. The extra-motor frontotemporal cortical regions are mildly affected. To the best of our knowledge, CHCHD10 p.R15L is the first fALS mutation to present predominantly as FAS.

### Neuropathology reveals greater CHCHD10 aggregation in cervical spinal cord

To determine if there is a neuropathologic correlate to these clinical findings, we examined the brain and spinal cord of one deceased member of the family. The individual’s clinical course was similar to other family members. Weakness first developed in the right arm when she was in her 50s, with spread to the contralateral arm approximately three years later. She subsequently developed bulbar symptoms with trouble swallowing and a soft voice, and a severe cough that precluded the use of pressure support ventilation. She had leg weakness eight years after symptom onset, and she died one year later.

Neuropathological assessment of her spinal cord showed more severe involvement of the cervical spinal cord correlating to her clinical symptoms; subtotal loss of motor neurons in the anterior horn was observed throughout the spinal cord with relative sparing of the lumbar segments. Consistent with greater lower motor neuron (LMN) than UMN involvement, no significant neuronal loss or gliosis was seen in the cortex. Cortical sections stained for beta-amyloid, phosphorylated tau (AT8), and alpha synuclein were negative. No aberrant TDP-43 or phosphorylated TDP-43 (S409/410) immunolabeling was seen in cortical and spinal cord sections. CHCHD10 immunolabeling, however, revealed abundant neuropil threads in cervical segments and occasional fibrillar aggregates in residual motor neurons (Fig. 3A – I). Lumbar sections show a relatively lower density of neuropil threads compared to cervical sections (Fig. 3B and C); occasional fibrillar aggregates and rare round "glassy" inclusions were seen in residual lumbar motor neurons (Fig. 3D and E). CHCHD10 immunolabeling of the motor cortex shows diffuse neuropil threads throughout all cortical layers and scattered fibrillar aggregates in cortical neurons (Fig. 3I). Rare CHCHD10 threads and occasional neuronal inclusions were noted in the cingulate cortex. Extremely rare CHCHD10 threads and no neuronal inclusions were seen in the frontal cortex. Together these findings suggest a close correspondence between CHCHD10 protein misfolding in the CNS and the clinical pattern observed in symptomatic *CHCHD10* p.R15L mutation carriers.

**Figure 3.**
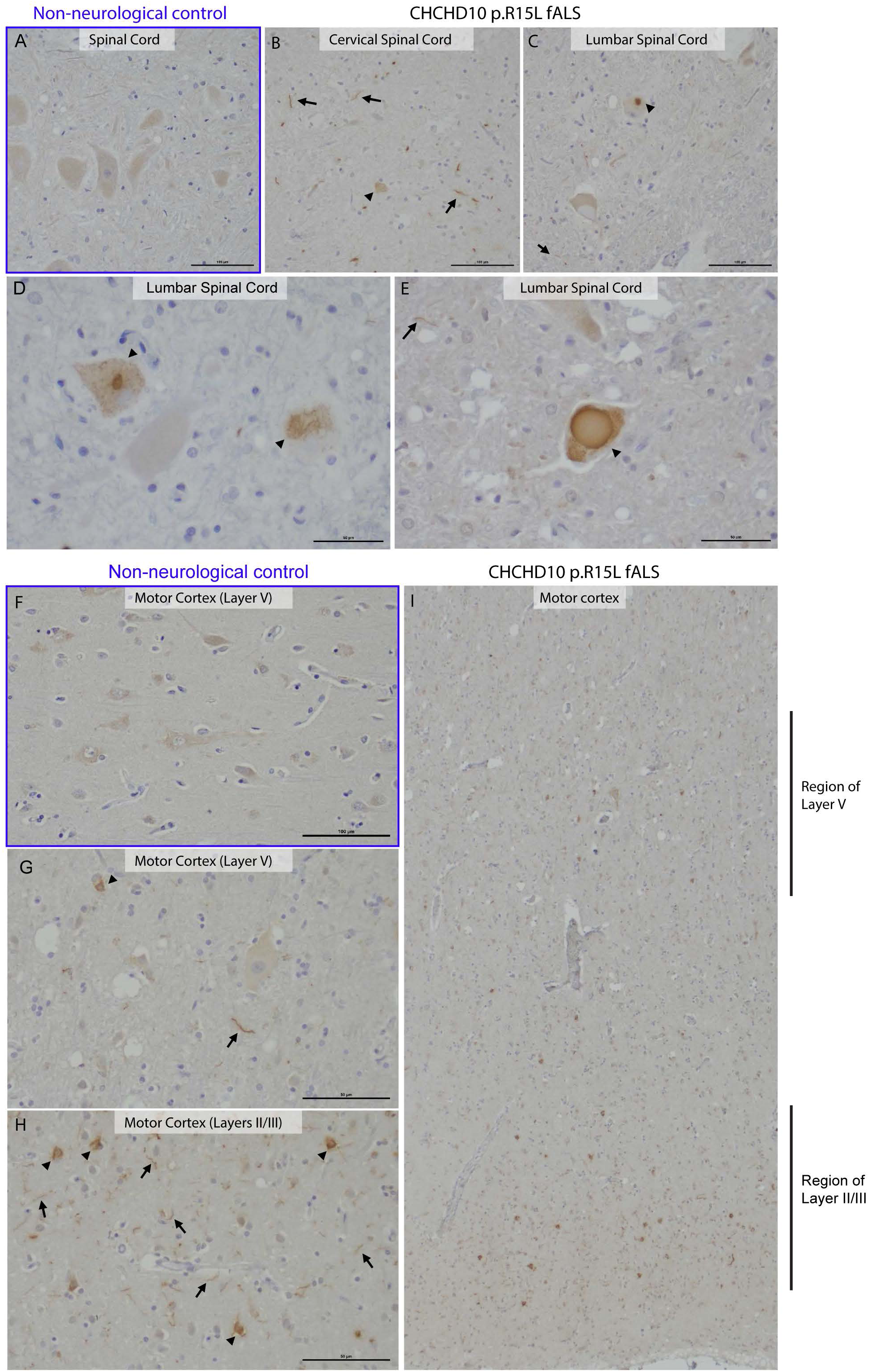
CHCHD10 pathology in spinal cord and motor cortex. (A) Spinal cord section from a non-neurological control showing weak diffuse cytoplasmic CHCHD10 immunohistochemistry. Scale bar 100 μm. (B) Cervical spinal cord showing scattered neuropil threads (arrows) and neuronal cytoplasmic inclusions in the soma of the residual motor neurons (arrowhead). Scale bar 100 μm. (C) Lumbar spinal cord showing rare threads (arrow) and neuronal cytoplasmic inclusions in the soma of the residual motor neurons (arrowhead). (D) High-power image of lumbar spinal cord highlighting fibrillar CHCHD10-positive aggregates in motor neurons (arrows). Scale bar 0 μm. (E) High-power image of lumbar spinal cord highlighting rare round and “glassy” CHCHD10-positive aggregates (arrowhead). Scale bar 100 μm. (F) High-power image of layer V of frontal cortex from a non-neurological control showing weak diffuse cytoplasmic staining in neurons. Scale bar 100 μm. (G) High-power image of layer V of motor cortex showing diffuse neuropil threads (arrow) and occasional neuronal cytoplasmic aggregates (arrow head). Scale bar 0 μm. (H) High-power image of layers II/III of motor cortex showing diffuse neuropil threads and neuronal cytoplasmic aggregates. Scale bar 0 μm. (I) Low-power image of motor cortex showing diffuse neuropil threads and neuronal cytoplasmic aggregates.

### Pathogenic CHCHD10 p.R15L variant retains partial protein expression and function in cell lines

We next considered the pathogenetic mechanism of the p.R15L variant. Prior studies using a transformed patient fibroblast line or transformed lymphoblasts concluded that CHCHD10 p.R15L is mostly unstable (at either the RNA or protein level), which was interpreted to favor a LoF model of pathogenesis ^23,24^. These studies had several limitations, however: in lymphoblasts, only mRNA expression was assessed ^24^, and, in fibroblasts only one patient cell line was available for comparison ^23^; the lines were compared to unrelated (i.e., non-isogenic) controls; the mutation was heterozygous and so expression of CHCHD10 from the disease-allele had to be inferred from the total CHCHD10 levels (WT + disease allele); and robust assays were not available at the time to assess CHCHD10 function.

This prompted us to revisit two questions: is the CHCHD10 p.R15L variant stable, and is it functional? For this purpose, we developed a more extensive set of primary fibroblast lines from affected and unaffected mutation carriers, isogenic iPSC cell lines with and without the mutation in homozygous state, and a knock-in (KI) transgenic mouse model harboring a humanized CHCHD10 p.R15L allele.

We first established primary fibroblast cell lines from twelve mutation carriers (five affected and seven unaffected with ALS). These were compared to primary dermal fibroblast lines from available healthy controls and an unrelated disease control group (idiopathic Parkinson’s disease). Notably, CHCHD10 protein levels were variable from line to line among both healthy and unrelated disease control lines, suggesting a high physiologic variance of CHCHD10 protein levels in primary dermal fibroblasts (Fig. 4A and B). CHCHD10 levels in p.R15L mutation carriers were lower on average but fell within the control range; the means were not significantly different between the two groups (Fig. 4B).

**Figure 4.**
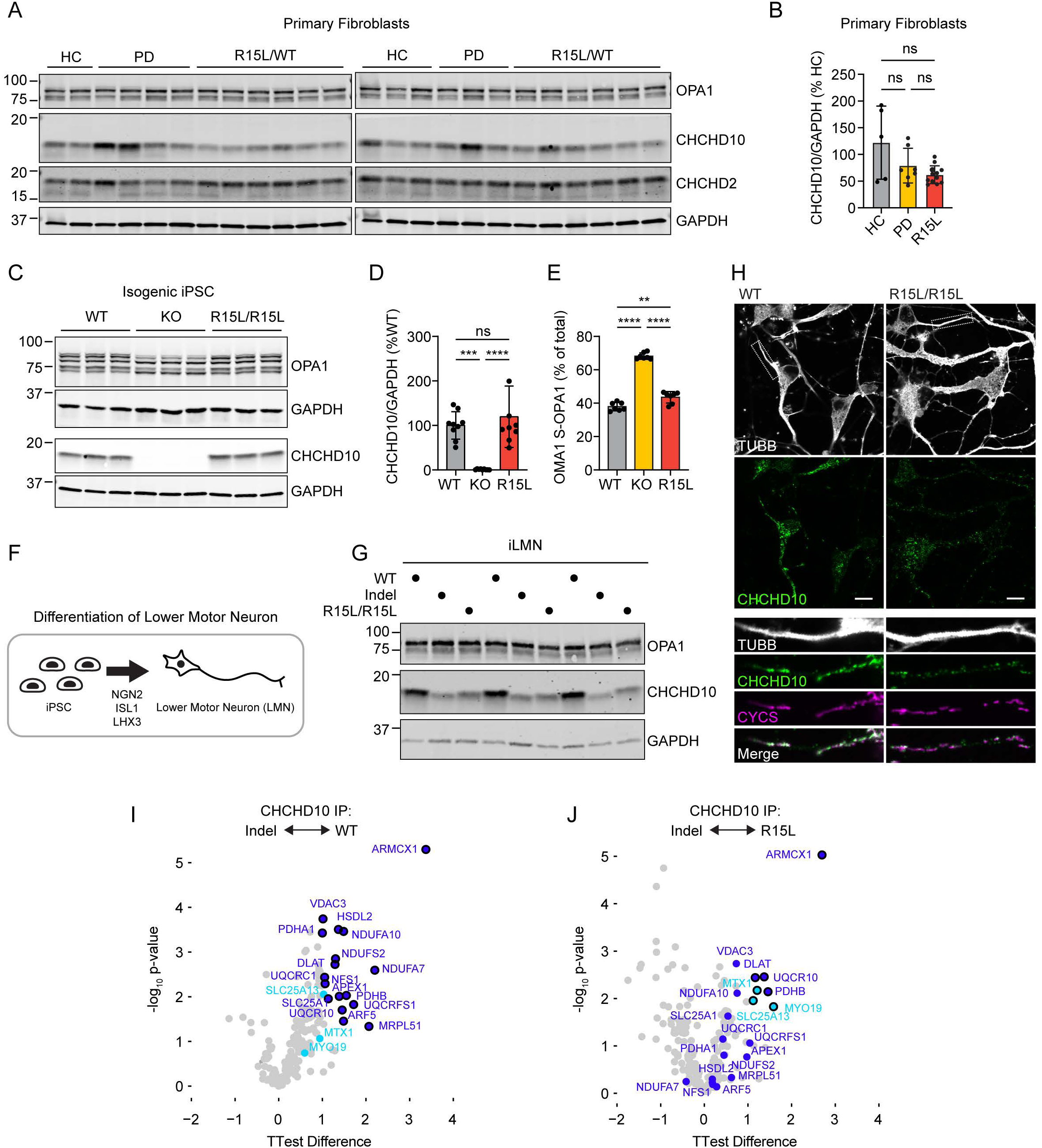
CHCHD10 R15L is stably expressed and partially functionally in human cells. (A and B) CHCHD10 protein levels were measured by immunoblotting from primary dermal fibroblast cell lines. Lines were established from 12 *CHCHD10* p.R15L mutation carriers (5 affected and 7 unaffected), 5 healthy controls, and 7 unrelated disease controls (with idiopathic Parkinson’s disease). (C - E) CHCHD10 protein levels (C and D) and OPA1 cleavage by OMA1 (C and E) were measured by immunoblotting for isogenic WT, *CHCHD10* KO, and *CHCHD10* R15/R15L iPSC cell lines. OMA1 activation is reflected by the cleavage of L-OPA1 (*a* and *b* isoforms) to specific S-OPA1 isoforms (*c* and *e* isoforms). Statistical analysis was performed using the Brown-Forsythe ANOVA test followed by Dunnett’s T3 multiple comparisons test. (F) Schematic demonstrates the method used to generate induced Lower Motor Neuron like cells (iLMNs) from iPSCs through the doxycycline induced expression of transcription factors *NGN2*, *ISL1*, and *LHX3*. (G) Immunoblotting of isogenic WT, *CHCHD10* indel, and *CHCHD10* R15/R15L iLMNs after 9 days *in vitro*. Data is representative of two separate differentiations. (H) Representative confocal image of CHCHD10 in iLMNs from isogenic WT and R15L/R15L iLMNs. Mitochondria are marked by cytochrome *c* (CYCS) and neuronal processes by β3-Tubulin (TUBB). Scale bars = 10 μm. Data is representative of N = 2 wells from a single differentiation. (I and J) Volcano plot of affinity purification mass spectrometry data following immunoprecipitation of endogenous *CHCHD10* from WT (left) and R15L/R15L (right) iPSC cells compared to CHCHD10 indel cells. Dark blue datapoints were significant interactors (FDR ≤ 0.05 and log2 fold change ≥ 1) of CHCHD10 in WT iPSC cells; light blue datapoints were significant interactors of CHCHD10 in R15L/R15L iPSC cells. Data is from N ≥ 3 wells of cells cultured and processed in parallel. In all panels, ns, *, **, ***, ***” correspond to not-significant, p ≤ 0.05, p ≤ 0.01, p ≤ 0.001, and p ≤ 0.0001, respectively. For (Fig. 3D an DE) an ordinary one-way ANOVA test followed by Dunnett’s multiple comparison’s test with a single pooled variance was performed, as the data was normally distributed with equal standard deviations.

To achieve better control over the genetic background, we next introduced the p.R15L variant in a homozygous state into the WTC induced pluripotent stem cell (iPSC) line (hereafter, R15L/R15L). For comparison, we generated a CHCHD10 depleted iPSC line by introducing compound heterozygous indel mutations (c.270-299del; c.288delA) in exon 2 (hereafter, indel), and a CHCHD10 KO line. CHCHD10 levels were similar in the parent and R15L/R15L iPSC cell lines and were decreased or absent in the indel and KO lines, respectively (Fig. 4C– E and S2A - C). As all CHCHD10 protein is either mutant or WT, the endogenous localization of the mutant can be readily determined by immunocytochemistry. We found that both WT and R15L CHCHD10 localized to mitochondria in a similar punctate pattern (Fig. S3). Some cytosolic and nuclear staining was determined to be background, as it was also present in the KO line (Fig. S3).

We next differentiated iPSCs to iLMNs through expression of the transcription factors *NGN2*, *ISL1*, and *LHX3*. All lines could be efficiently differentiated into iLMNs apart from the KO line, which was excluded from this analysis (Fig. S2D and data not shown). The overall morphology of the iLMNs was similar for the WT and R15L/R15L lines. Interestingly, CHCHD10 protein levels decreased in the R15L/R15L line in comparison to the parent line upon differentiation (Fig. 4G - H). This was observed both by immunoblotting and immunocytochemistry. Even in iLMNs, however, substantial CHCHD10 p.R15L protein was present within mitochondria (Fig. 4G). Together, these findings suggest that CHCHD10 p.R15L protein is stable in rapidly dividing iPSC cells but is partially destabilized (or fails to upregulate) upon differentiation into post-mitotic iLMNs.

Next, we examined whether endogenous CHCHD10 R15L can substitute for WT CHCHD10 in iPSC cells. We previously determined that loss of both CHCHD10 and its paralog CHCHD2 strongly activates a mitochondrial stress response initiated by the peptidase OMA1 ^13,43^. OMA1 activity can be tracked by examining its cleavage of the mitochondrial protein L-OPA1 (isoforms *a* and *b*) to S-OPA1 (isoforms *c* and *e*). As CHCHD2 expression was absent in our parent and mutant iPSC lines (as observed in many established pluripotent stem cell lines ^44–46^), we reasoned that loss of CHCHD10 activity should trigger the OMA1 stress response. As expected, the OMA1 stress response was strongly activated in the CHCHD10 indel and KO cell lines, consistent with substantial loss of CHCHD10 function in these two lines (Fig. 4C and E and S2C). By contrast, only weak OMA1 activation was observed in the CHCHD10 R15L/R15L line (Fig. 4C and E and S2C). This demonstrates that CHCHD10 R15L can partially substitute for WT CHCHD10.

The function of CHCHD2 and CHCHD10 is thought to depend on their transient interactions with other proteins within intermembrane space and IMM of mitochondria. To compare the protein-protein interactions (PPIs) among variants, we first established PPIs for WT CHCHD10 by affinity purification mass spectrometry (AP-MS). For this purpose, we used an antibody that was specific for CHCHD2 and CHCHD10 orthologs; the indel line (depleted of CHCHD10) was used a negative control for comparison. This revealed several PPIs of WT CHCHD10 including several subunits of respiratory complexes I and III (i.e., NDUFA10, NDUFA7, NDUFS2, UQCRFS1, UQCRC1, and UQCR10) (Fig. 4I and J and Table S2). Consistent with retained function of the R15L variant, we found that endogenous CHCHD10 R15L had higher affinity for WT PPIs relative to the indel line (apart from one protein, NDUFA7) (Fig. 4I and J and Table S2). Overall, however, these interactions were weaker for R15L than WT. Together, these findings establish that the R15L variant retains substantial expression and function, with a phenotype that is intermediate between the indel line and the WT line.

### Pathogenic CHCHD10 p.R15L variant retains partial protein expression and function in knock-in mice

Next, we tested the ability of *CHCHD10* p.R15L to substitute for *Chchd10* WT *in vivo*. For this purpose, we generated a humanized CHCHD10 p.R15L knock-in (KI) mouse, in which the coding and intronic regions of mouse *Chchd10* gene were replaced with the sequence of the patient *CHCHD10* p.R15L allele (hereafter, *Chchd10^hR15L^*) (Fig. 5A). This resulted in expression of human CHCHD10 p.R15L protein from the endogenous mouse *Chchd10* promoter. The mice were analyzed on a mixed C57BL/6 and SJL background. This KI model contrasts with a previously reported mouse models in which human CHCHD10 p.R15L was over-expressed from an exogenous promoter ^25,26^. *Chchd10^hR15L/+^* mice appeared grossly normal with a similar body weight as their littermates beyond one year of life (Fig. 5B). While a more detailed characterization of this mouse line is planned for future work, here, we sought to establish whether human CHCHD10 p.R15L is expressed in mature neurons in the brain and spinal cord and whether *Chchd10^hR15L^* can functionally replace WT mouse *Chchd10 in vivo*.

**Figure 5.**
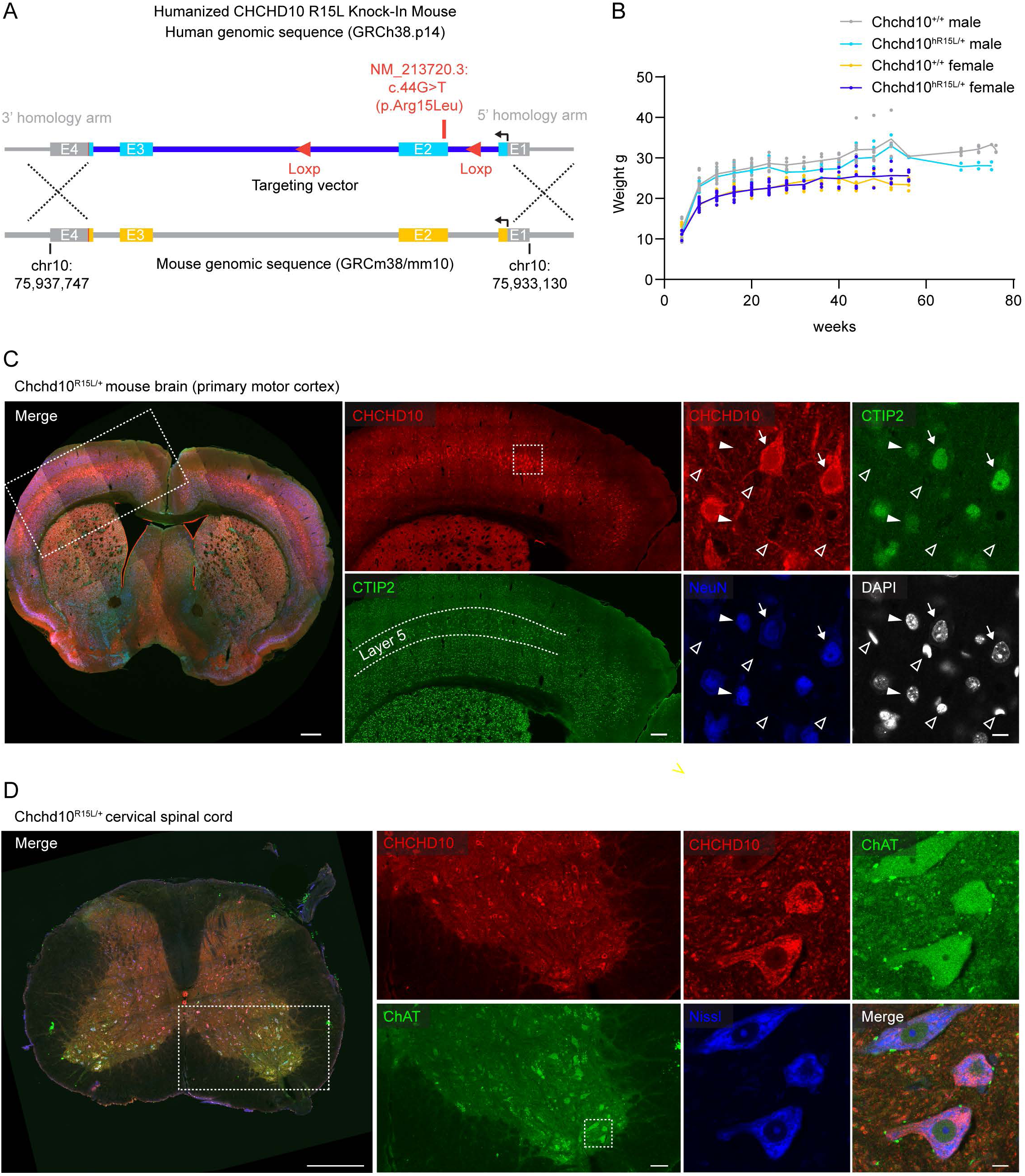
Expression of CHCHD10 R15L in brain and spinal cord of humanized knock-in mouse. (A) Schematic demonstrating the generation of the Chchd10^hR15L/+^ knock-in (KI) mouse line. The coding region of human CHCHD10 inclusive of introns was knocked into the endogenous mouse *Chchd10* locus. The haplotype was identical to that of the USALS#5 family described in (Fig. 1). Loxp sites were added to the introns flanking exon 2. (B) Body weights of Chchd10^hR15L/+^ knock-in (KI) and WT littermates. Chchd10^hR15L/+^ mice were grossly normal up to 1 year. (C) Immunofluorescence of a coronal brain section at the level of the primary motor cortex from Chchd10^hR15L/+^ mice. Within the cortex CHCHD10 immunostaining (red) was substantially higher in layer 5, identifiable by CTIP2 immunostaining (green). This layer contains the extratelecephalic upper motor neurons cells that project to the spinal cord through the corticospinal track. Within layer 5, CHCHD10 immunostaining was highest in CTIP2 bright neurons and lowest in non-neuronal cells (DAPI+ but NeuN-). Dotted rectangles in images indicate the approximate areas of the magnified images that appear immediately to the right. Neurons with high CTIP2 expression are indicated by arrows. Those with lower CTIP2 expression are indicated by closed arrow heads. DAPI positive nuclei that were negative for the neuronal nuclear marker NeuN are indicated by open arrowheads. Scale bars are 500 μm (left image), 200 μm (middle images), and 10 μm (right images), respectively. (D) Immunofluorescence of a coronal spinal cord section at the level of the cervical enlargement from Chchd10^hR15L/+^ mice. CHCHD10 (red) immunostaining is strongest in the ChAT (green) positive lower motor neurons. Dotted rectangles in images indicate the approximate areas of the magnified images that appear immediately to the right. Scale bars are 500 μm (left and middle images) and 10 μm (right images), respectively.

Comparison of brain from 60-week-old mice showed higher CHCHD10 reactivity in *Chchd10^R15L/+^* vs. *Chchd10^+/+^* mice (Fig. S4A). This is likely due to the higher affinity of the antibody (raised against human CHCHD10) for human vs. mouse CHCHD10, as was also apparent by immunoblotting (below). Notably, in the primary motor cortex, CHCHD10 was strongly enriched in large pyramidal cells in layer 5, identified by greater CTIP2 immunostaining (Fig. 5C and S4B). This population includes the upper motor neurons that are primarily affected in ALS. CHCHD10 was observed in a pattern suggestive of mitochondrial localization for both WT and p.R15L variants. Aggregates of CHCHD10 were not observed. In layer 5, CHCHD10 staining was lower in NeuN positive neurons with low CTIP2 staining (Fig. 5C, closed arrow heads) and around non-neuronal nuclei identified by DAPI+/NeuN-staining (Fig. 5C, open arrow heads). CHCHD10 R15L expression could also be visualized in the cervical spinal cord, where its expression was highest in ChAT positive lower motor neurons (Fig. 5D). This demonstrates strong expression of human CHCHD10 p.R15L driven from the endogenous mouse promoter in upper and lower motor neurons *in vivo*.

We next sought to determine whether the *Chchd10^hR15L^*allele can substitute for WT mouse *Chchd10* and *Chchd2* and thus retains function *in vivo*. For this purpose, mice with this allele were crossed onto a *Chchd2*^-/-^; *Chchd10*^-/-^ background to test if the allele can suppress the strong OMA1-DELE1 mitochondrial integrated stress response (mt-ISR) triggered by loss of both CHCHD2 and CHCHD10 ^13,43^ (Fig. 6A and B). This stress response is particularly strongly activated in the heart muscle and so we examined heart as well as spinal cord lysates.

**Figure 6.**
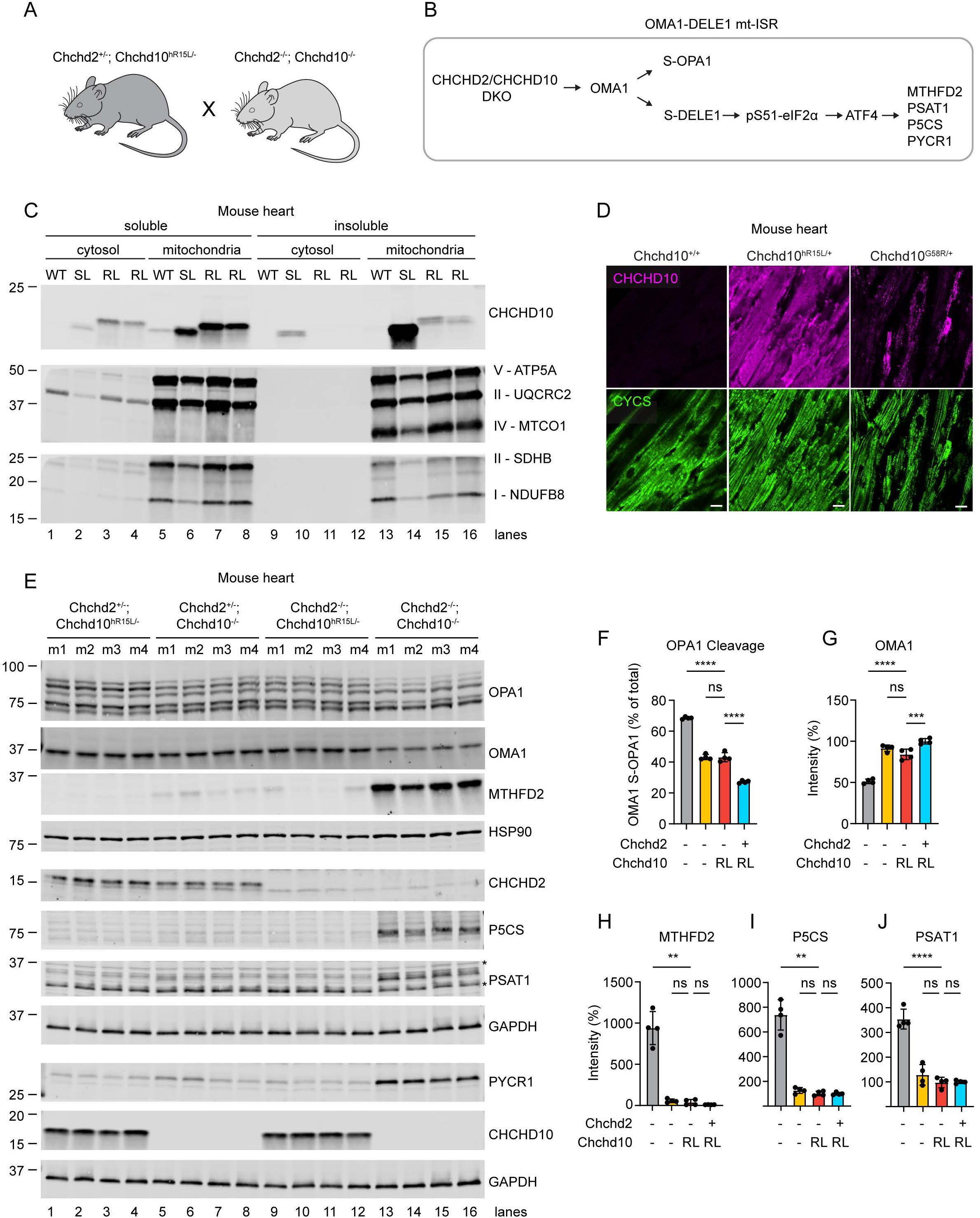
CHCHD10 R15L is functional *in vivo*. (A) Schematic shows the cross used in this set of experiments to test whether a single CHCHD10 R15L allele can suppress the mitochondrial integrated stress response (mt-ISR) activation observed in the CHCHD2/CHCHD10 double knockout mouse. This cross produces the four genotypes that are compared in (Fig. 5E). (B) Schematic shows activation of the OMA1-DELE1 mt-ISR. Activation of the mt-ISR is evident both in the cleavage of L-OPA1 by OMA1 and the upregulation of enzymes that are targets of the transcription factor ATF4. These targets include MTHFD2, PSAT1, P5CS, and PYCR1. (C) Solubility and localization of CHCHD10 was determined for *Chchd10^+/+^; Chchd2^+/+^* (WT) and *Chchd10^hR15L/-^; Chchd2^-/-^*(RL) by immunoblotting following subcellular fractionation of heart tissues. Heart tissue from a *Chchd10^S59L/+^; Chchd2^+/+^* (SL) mouse was additionally examined as a positive control. Heart tissues were fractionated by centrifugation into a cytosolic fraction and a mitochondria-enriched heavy membrane fraction. These fractions were further fractionated by differential detergent solubilization into soluble and insoluble fractions. CHCHD10 R15L was predominately in the soluble mitochondrial fraction. As expected, OXPHOS subunits for complex I - V were found predominately in the mitochondrial fraction. OXPHOS subunit expression was decreased in the SL mouse sample, as expected, but was not decreased in samples from RL mice relative to WT. These data are representative of 2 WT, 4 RL, and 2 SL mice. RL mice were approximately 8 weeks of age. WT and SL mice were approximately 40 weeks old. (D) Immunofluorescence of mouse heart tissue. Mitochondria are indicated by cytochrome *c* (CYCS) immunostaining. Stronger signal is observed for *Chchd10^hR15L/+^*plausibly because of the higher affinity of the antibody for human CHCHD10. The distribution of CHCHD10, however is similar to *Chchd10^+/+^* mice and distinct from CHCHD10 aggregation observed in *Chchd10^G58R/+^* mice. (E - J) Immunoblotting of heart lysates from mice produced by the cross shown in (Fig. 5A). Suppression of OMA1 activation by a single *Chchd10* hR15L allele is evident in the increased L-OPA1/S-OPA1 ratio (E and F) and restoration of OMA1 levels (which decrease on OMA1 activation) (E and G). Downstream of OMA1, enzymes that are transcriptionally upregulated by the mt-ISR are likewise normalized by expression of a single *Chchd10* hR15L allele (E and H – J). Mice were approximately 8 weeks of age. In all panels, ns, *, **, ***, ***” correspond to not-significant, p ≤ 0.05, p ≤ 0.01, p ≤ 0.001, and p ≤ 0.0001, respectively. For (Fig. 5F, G, and J) an ordinary one-way ANOVA test followed by Dunnett’s multiple comparison’s test with a single pooled variance was performed, as the data was normally distributed with equal standard deviations. For (Fig. 5H and I) a Brown-Forsythe and Welch ANOVA test followed by a Dunnett’s T3 multiple comparisons test was performed as the standard deviations were not equal.

We first considered the localization and solubility of CHCHD10 p.R15L. Human CHCHD10 p.R15L was readily observed in both the heart and spinal cord lysates (Fig. 6C and S4D). It migrated at a slightly higher apparent molecular weight than WT mouse CHCHD10, consistent with its larger size (142 AA vs. 138 AA) and decreased net charge. The band for human CHCHD10 was also more intense than that for its mouse counterpart, consistent with our observations by immunofluorescence above. Similar to WT, CHCHD10 p.R15L localized to the mitochondrial fraction in hearts from *Chchd10^hR15L/-^; Chchd2^-/-^* mice (Fig. 6C, lane 6, 7, and 8). It was also soluble in mild detergent (1% Triton) (Fig. 6C, lane 15 - 16 vs. 7 - 8). This was in contrast to the ALS/FTD variant CHCHD10 p.S59L, which was found predominately in the insoluble fraction, as previously reported (lane 14 vs. 6) ^13,14^. Notably, there was no reduction in OXPHOS subunits in the *Chchd10^hR15L/-^; Chchd2^-/-^* mouse hearts (lanes 7 and 8 vs. 5), in contrast to a moderate reduction in the *Chchd10^S59L/+^* mouse (lanes 6 vs. 5), as reported previously ^14,21^. This is consistent with the absence of a gross phenotype at 1 year of age. Similarly, we found no CHCHD10 protein aggregates in the heart of a *Chchd10^hR15L/+^* mouse (Fig. 6D), in contrast to what has been observed previously for *Chchd10^S59L/+^* and *Chchd10^G58R/+^* ^13,14^. As expected, CHCHD10 signal was higher in the *Chchd10^S59L/+^* heart, likely reflecting the higher affinity of the antibody for human CHCHD10.

We next assessed the ability of the CHCHD10 p.R15L to suppress the OMA1-DELE1 mt-ISR that is activated in the absence of CHCHD2 and CHCDH10 ^20,43^. Strikingly, a single *Chchd10^hR15L^*allele normalized OMA1 activation (restoration of OPA1 cleavage and OMA1 self-cleavage) as well as the enzymes that are transcriptionally upregulated by the mt-ISR downstream of ATF4 ^43^ (Fig. 6E - J). Notably, the degree of rescue was similar to that conferred by a single WT *Chchd2* allele. The combination of a single *Chchd2* allele and a single *Chchd10^hR15L^* was additive, resulting in further suppression of OMA1 activation (Fig. 6F). Thus, *Chchd10^hR15L^* can substitute at least in part for WT *Chchd2* and *Chchd10 in vivo*. Together these genetic findings suggest that CHCHD10 R15L is a partially functional allele with substantial activity both *in vivo* and in cultured cells.

### Heterozygous CHCHD10 loss of function variants are not a highly penetrant cause of ALS in contrast to p.R15L

Our findings in model systems suggest that CHCHD10 p.R15L can largely replace WT CHCHD10. This demonstrates that the p.R15L variant is a hypomorph (i.e., has reduced function) rather than a LoF variant. We next considered whether carrying a LoF CHCHD10 allele is sufficient to cause a highly penetrant form of ALS, as would be predicted by the LoF model of p.R15L pathogenesis. The UKBB, a large cohort study, has recently been used to establish the penetrance of repeat expansions in *C9orf72*, the most common genetic cause of ALS ^47^. In this study, 10.3% of C9orf72 mutation carriers were found to have ALS ^47^. We reasoned that if haploinsufficiency of CHCHD10 is a cause of highly penetrant fALS then a similar penetrance should be observed for heterozygous LoF mutations in *CHCHD10* in the UKBB. Analyzing the whole-exome sequencing data in the UKBB, we identified 7 predicted LoF variants in *CHCHD10* in 31 individuals, all occurring in the heterozygous state (Table S3). These LoF variants were more common than the ultrarare variants shown to segregate with disorders of motor neurons (p.R15L, p.S59L, and p.G66V), which were not present in the UKBB. Among the LoF variants, one was predicted to disrupt a canonical splice site. The others were nonsense or frameshift variants that would result in loss of conserved cysteine residues required for CHCHD10 localization and function. Notably, none of the participants carrying these LoF variants (0/31) had been diagnosed with ALS. This suggests that heterozygous LoF variants in *CHCHD10* are not a penetrant cause of fALS, in contrast to p.R15L.

### CSF proteomics reveal accumulation of CHCHD10 in p.R15L variant carriers

We next evaluated CHCHD10 protein levels in cerebrospinal fluid (CSF) of p.R15L carriers. We reasoned that CHCHD10 levels would be lower if the p.R15L variant substantially destabilizes CHCHD10 *in vivo*. By contrast, CHCHD10 levels may be higher if the variant causes the protein to misfold in a manner that hinders its clearance, as in a toxic GoF model.

To test these predictions, we first measured CHCHD10 levels using the Olink proteomics platform. Strikingly, CHCHD10 levels were approximately two-fold higher in both symptomatic and presymptomatic R15L carriers compared to healthy volunteers (Fig. 7A – C and Table S4). Indeed, CHCHD10 was the most significant differentially regulated mitochondrial protein for both symptomatic and presymptomatic mutation carriers (of 232 mitochondrial proteins), and the fourth most significantly differentially regulated protein (DEP) overall for symptomatic mutation carriers. Structurally related mitochondrial proteins, including the paralog CHCHD2, were not substantially changed, demonstrating that the elevation is highly specific for CHCHD10. Interestingly, CHCHD10 levels were also slightly elevated in sporadic ALS and C9orf72 ALS groups, reaching statistical significance for the latter group (Fig. 7C). This raises the possibility that CHCHD10 may mark and/or contribute to other forms of ALS. These findings were confirmed in the independent analysis using the SomaScan proteomics platform (Table S5). CHCHD10 was significantly elevated in both presymptomatic and symptomatic CHCHD10 R15L carriers relative to controls (Fig. 7D and S5).

**Figure 7.**
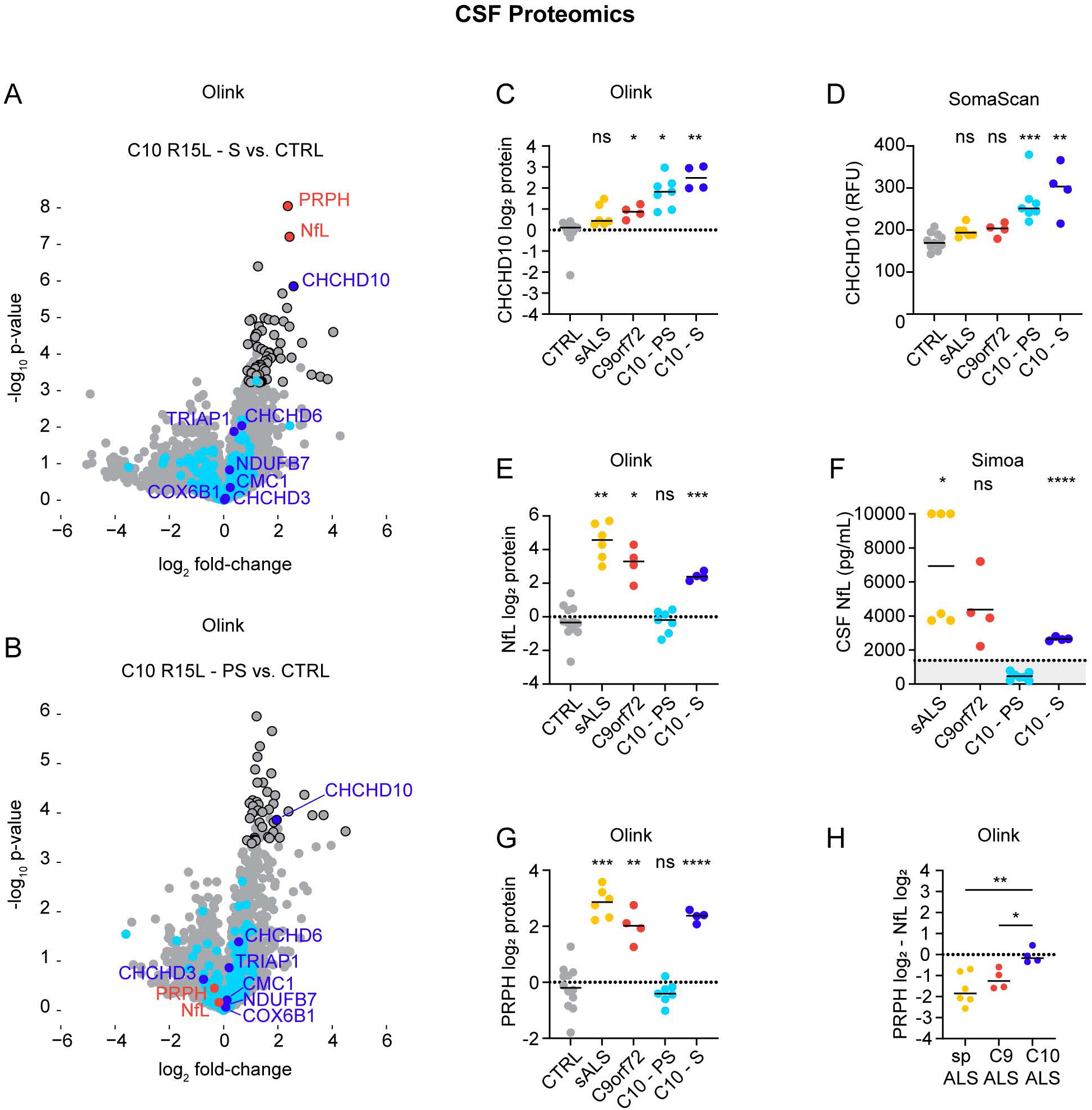
CSF proteomics reveal CHCHD10 and NfL elevation in CHCHD10 R15L ALS. (A - B) Volcano plots of CSF proteomics with the Olink Explore HT platform comparing symptomatic (A) and presymptomatic (B) CHCHD10 R15L mutation carriers to healthy controls. Uncorrected p-value is shown on the y-axis. Significant differentially expressed proteins (FDR < 5% in the one-way ANOVA across all groups and the t-test for the comparison of interest) are outlined in black. Mitochondrial proteins are blue with CHCH-domain containing proteins shown in dark blue and others in light blue. The neurofilaments NfL and PRPH are shown in red. (C) Dot plot of CHCHD10 protein levels for individuals measured by Olink proteomics in the same experiment depicted in (A – B). (D) Dot plot of CHCHD10 protein levels for individuals measured by Somascan proteomics in the same experiment depicted in (Fig. S5). (E) Dot plot of NfL protein levels for individuals measured by Olink proteomics in the same experiment depicted in (A – B). (E) Dot plot of NfL protein levels for individuals measured using a SIMOA assay. Dotted line indicates the cutoff value of 1431 pg/mL previously suggested for ALS ^49^. CHCHD10 presymptomatic (C10-PS) was used as the control for pairwise statistical comparisons. (G) Dot plot of PRPH protein levels for individuals measured by Olink proteomics in the same experiment depicted in (A – B). (H) Dot plot shows the ratio of PRPH to NfL protein levels for the indicated groups. Adjusted p-values are shown for (C, D, E, and G) to reflect multiple comparisons across proteins in the source dataset. P-values for (F and H) are adjusted for the multiple comparisons shown in graphs. In all panels, ns, *, **, ***, ***” correspond to not-significant, p ≤ 0.05, p ≤ 0.01, p ≤ 0.001, and p ≤ 0.0001, respectively.

Together these results establish that CHCHD10 levels are elevated approximately two-fold in the CSF of p.R15L mutation carriers, consistent with a toxic GoF model of pathogenesis. They also demonstrate for the first time that CHCHD10 levels can be measured in the CSF of patients with neurodegeneration related to mutations in CHCHD10. Importantly, this suggests that CSF CHCHD10 levels may be tracked as a biomarker for CNS target engagement in therapeutic trials.

### Neurofilament light chain and peripherin differentiate symptomatic and presymptomatic p.R15L carriers

We next considered which CSF biomarkers differentiate symptomatic from presymptomatic p.R15L mutation carriers. The top two DEPs for this comparison in the Olink data were the neuronal cytoskeletal proteins peripherin (PRPH) and neurofilament light chain (NfL) (Fig. 7A, B, E, and G and Table S4). NfL is expressed in neurons in both the CNS and the peripheral nervous system (PNS), whereas PRPH is restricted to the PNS and lower motor neurons ^48^. NfL was also the top DEP in the SomaScan analysis (PRPH was not measured) (Figure S5 and Table S5).

To obtain absolute quantification of NfL we independently analyzed CSF samples using a Simoa-based ELISA (Fig. 7F). This confirmed that NfL was above the proposed ALS cutoff value for all affected CHCHD10 R15L participants ^49^, and below the value for all unaffected CHCHD10 R15L participants. Among the unaffected participants was carrier in her 70s with younger affected siblings. This provides biological evidence for a discrete onset of neurodegeneration in CHCHD10 R15L fALS. It further suggests that NfL levels may be useful to monitor for phenoconversion among mutation carriers.

As PRPH is expressed only in LMN but not UMN, we reasoned that the ratio of PRPH to NfL may reflect the degree of LMN vs. UMN degeneration. Consistent with our clinical observations, the PRPH/NfL ratio was higher in CHCHD10 R15L ALS than C9orf72 or sALS (Fig. 7H). This provides further biological evidence that CHCHD10 p.R15L causes a distinct form of fALS, characterized by preferential degeneration of lower motor neurons.

Together, our proteomic analysis of CSF identifies biomarkers for both the trait and the state of CHCHD10 p.R15L ALS. These may be used to track phenoconversion as well as target engagement and treatment response in future gene-directed therapeutic trials.

## Discussion

In a comprehensive clinical, neuropathological, and biochemical characterization of a large family, we establish that the CHCHD10 p.R15L variant causes FAS, likely through a GoF mechanism. We additionally identify CSF biomarkers that may be used to assess phenoconversion, target engagement, and treatment response.

Our finding that the *CHCHD10* p.R15L variant presents almost exclusively as FAS is consistent with more limited reports of two German families and one sporadic Canadian patient with the variant ^8,10^. FAS is an uncommon ALS variant, occurring in about 5.5% patients ^1^. It has been reported to be enriched in some fALS cases caused by mutations in *hnRNPA1* (particularly, the p.P340S variant), *NEK1*, and *TARDBP* ^50–54^. However, in these genetic forms of fALS there is greater phenotypic heterogeneity than we found is the case with *CHCHD10* p.R15L. Thus, clinicians should think of the *CHCHD10* p.R15L variant, when ALS presents as FAS in the setting of a family history. These findings also have implications for the design of gene-directed therapy trials. Motor function measures focused on the upper extremities, but not lower extremities, will be most useful for tracking treatment response. Additionally, these findings suggest a strategy for early detection of phenoconversion; a simple finger tapping task readily distinguished presymptomatic and symptomatic mutation carriers. A remote version of this task could allow monitoring of symptom onset to guide treatment initiation. While motor features dominate, the ALS specific cognitive screening tool, ALS-CBS, was abnormal in 80 percent of the symptomatic and 22 percent of the presymptomatic participants, suggesting some degree of cognitive change. Furthermore, detailed neuropsychological testing showed a pattern of mild executive dysfunction consistent with reports of ALS associated cognitive change and indicative of a broader neurodegenerative process ^55^.

This stereotyped FAS presentation also distinguishes *CHCHD10* p.R15L from other variants in *CHCHD10* causing motor neuron disease – in particular, p.S59L and p.G66V. In contrast to p.R15L, p.S59L patients most often presented with bulbar symptoms and/or ataxia, with most also having cognitive dysfunction and mitochondrial myopathy ^7^. The p.G66V variant is also stereotyped in its presentation but typically affects the lower extremities first ^11^, in stark contrast to the upper extremity predominance of the p.R15L variant. Notably, this suggests that each of the three variants in *CHCHD10* has a different predilection for LMNs in the bulbar, cervical, and lumbar regions. This pattern is unlikely to reflect a gradient of CHCHD10 function. Rather, it suggests that each variant is preferentially toxic to a different subset of motor neurons. We hypothesize that the molecular basis for this genotype-phenotype variation may be different misfolded conformations of CHCHD10 resulting from each mutation. In support of this mechanism, two *CHCHD10* variants p.G58R and p.S59L with differential toxicities in myocytes and neurons were recently shown to form aggregates with differential solubility and aggregate morphology *in vivo* ^13,14^. Additionally, CHCHD10 was recently shown to form amyloids, *in vitro* and in heart muscle *in vivo*, with different variants adopting distinct folds ^56^. Also, consistent with this view, we observed CHCHD10 aggregates in the brain and spinal cord that closely matched the clinical phenotype of *CHCHD10* p.R15L ALS: LMNs were more severely affected than UMNs and the cervical region was more severely affected than the lumbar region. Notably, TDP-43 pathology was absent in both the brain and the spinal cord. The key findings of CHCHD10 aggregates and lack of TDP-43 pathology closely mirrors a neuropathological report from an unrelated Candidan patient with the CHCHD10 p.R15L variant ^16^. Importantly, the p.R15L mutation occurred on to two different *CHCHD10* haplotypes in these two neuropathological cases, reinforcing that the variant is the driver of the pathology. Collectively, these two neuropathologic cases strongly suggest that CHCHD10 p.R15L ALS does not cause a TDP-43 proteinopathy – as has been suggested by some transgenic mouse studies (for instance, ^26^) – but is strongly associated with accumulation of CHCHD10 protein aggregates.

This proposed mechanism implies the *CHCHD10* p.R15L variant is toxic GoF. However, some have argued for an haploinsufficiency model, instead, pointing to decreased expression of *CHCHD10* at the mRNA (in patient lymphoblasts) or the protein (in a patient fibroblast line) levels in patient lines heterozygous for the p.R15L variant ^23,24^. Here, we evaluated these assumptions, using novel patient and iPSC cell lines and a humanized KI mouse. While we find that *CHCHD10* p.R15L has decreased protein expression in some cellular contexts such as iLMNs, we demonstrate that *CHCHD10* p.R15L can largely substitute for WT *CHCHD10* in cultured cells and *in vivo*. Additionally, we demonstrate that CHCHD10 p.R15L is expressed in lower and upper motor neurons in a KI mouse model *in vivo* and is elevated in the CSF of mutation carriers. It is unclear at present why CHCHD10 p.R15L levels are lower in iLMNs than iPSCs, but cell context has also been shown to influence protein levels of other *CHCHD10* variants, such as p.G66V and p.G58R ^13,15^. Perhaps most striking, we find that a single human *CHCHD10* R15L allele nearly suppresses the strong OMA1-DELE1 mt-ISR in striated muscle of *Chchd2*/*Chchd10* DKO mice. This demonstrates that a heterozygous *CHCHD10* p.R15L allele while hypomorphic is not LoF. By contrast, we find that heterozygous LoF variants in CHCHD10, while more common than the ultrarare p.R15L variant, are not associated with highly penetrant forms of ALS in the population. Additionally, we show that p.R15L is a mutational hotspot, suggesting that this specific substitution (and not many others predicted to disrupt CHCHD10 stability and function) greatly increases the likelihood of developing ALS. Finally, we establish that CHCHD10 protein is markedly increased in the CSF of mutation carriers and forms aggregates in the neurites and soma of motor neurons. This would be unexpected if the p.R15L were completely destabilizing as suggested previously but is consistent with the view that the p.R15L variant may adopt an alternative fold that can both decrease its rate of CSF clearance and cause its aggregation in neurons. Notably, CSF elevation of CHCHD10 was observed in both presymptomatic and symptomatic p.R15L carriers. This argues against increased CHCHD10 release solely form degeneration of motor neurons, as other markers of neurodegeneration like NfL and PRPH were only elevated in the symptomatic mutation carriers. Rather, CHCHD10 may be released from cells of the CNS as part of quality control processes (such as, the exocytosis of autophagosomes/lysosomes) that maintain CNS homeostasis. Together, our results argue strongly for a toxic GoF model, similar to what has been established for the myopathy causing mutations in CHCHD10 ^13,14,21^.

Importantly, these findings suggest that *CHCHD10* p.R15L ALS may be amenable to gene directed therapy aimed at lowering levels of toxic CHCHD10 protein. Perhaps most promising is intrathecal delivery of antisense oligonucleotides (ASOs), as these have been successful for other genetic forms of fALS including those caused by mutations in SOD1 and FUS ^3,4^. As *CHCHD10* is likely inessential in mouse (and likely other mammals) ^18,20^, a nonallele-specific approach may be well tolerated and would have the added advantage of being applicable to other disease-causing variants in *CHCHD10*. Our analysis of human *CHCHD10* LoF mutation carriers in the UKBB further supports the safety of lowering CHCHD10 levels therapeutically.

To evaluate the effectiveness of these therapies, fluid biomarkers are needed to assess target engagement and treatment response, as shown by the recently successful trial of Tofersen, the FDA approved ASO against SOD1, and, similarly, in a recent trial of an ASO targeting FUS ^3,5^. Toward this end, we established that CSF levels of CHCHD10 and NfL are elevated in affected mutation carriers, suggesting that they may be used to track target engagement and treatment response, respectively. NfL and PRPH may also be useful for tracking phenoconversion of presymptomatic mutation carriers to guide treatment initiation. In conclusion, by establishing the disease mechanism and fluid biomarkers for CHCHD10 p.R15L fALS, our study helps set the stage for gene-directed therapeutic trials for a devasting form of fALS.

## Limitations

Our study has some important limitations. The design was cross-sectional, limiting inferences on the rate of progression. Future longitudinal studies will help refine the natural history of the disease, providing historical control data for therapeutic trials. Additionally, while our cell and mouse modeling approaches allow us to rule out a haploinsufficiency model of pathogenesis, these models have not yet yielded direct evidence for GoF. As KI mouse models of ALS often have subtle (if any) motor neuron involvement ^57^, a full characterization will require longer term study with a large cohort of Chchd10^R15L/+^ mice to determine if a motor neuron phenotype may develop at an advanced age.

## Data Availability

All data produced in the present study are available upon reasonable request to the authors

## Acknowledgments

This work utilized the computational resources of the NIH HPC Biowulf cluster (https://hpc.nih.gov). We thank Eric Shoubridge for helpful discussions and sharing critical material. We thank Prithviraj Rajebhosale for advice about upper and lower motor neuron markers for immunofluorescence of mouse tissue sections. We thank Yan Li and the NINDS Proteomics core for assistance MS-based proteomics. We thank Lijin Dong and the NEI Transgenic Core for assistance generating the Chchd10^R15L/+^ KI mouse. We thank Jizhong Zou and the NHLBI iPSC Core for assistance generating the CHCHD10 R15L/R15L and KO iPSC lines. We thank all of the families for their participation in this study.

## Supplementary material

Supplementary material is available at Brain online.

## Supplemental Figure Legends

**Figure S1.**
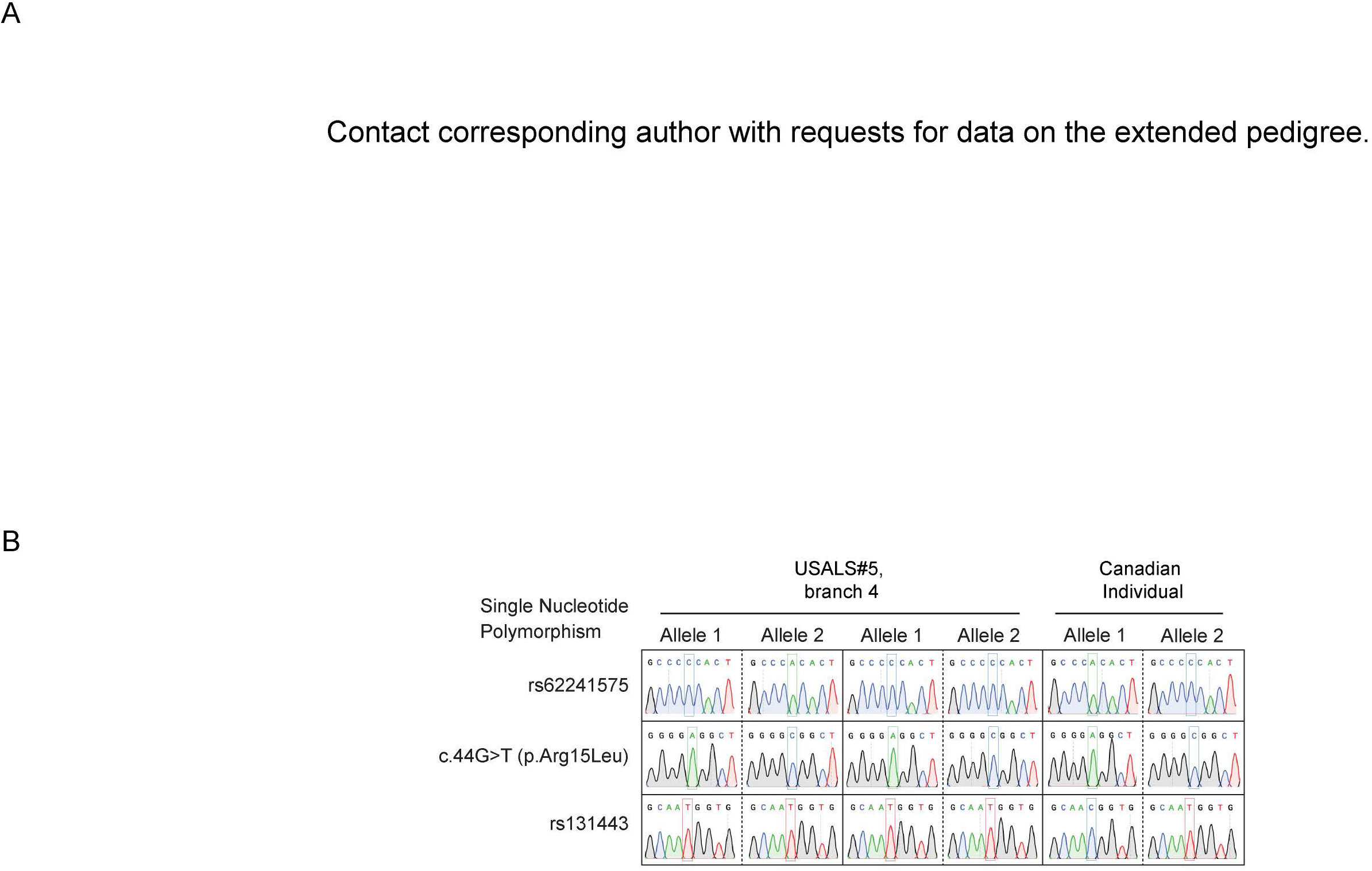
Extended pedigree from CHCHD10 R15L family. (A) Contact corresponding author with requests for data on the extended pedigree. (B) Sanger sequencing of cloned CHCHD10 alleles from indicated family members. Representative sequencing traces are shown across the pathogenic SNP and two SNPs that differed between the haplotype in the USALS#5 family and a previously reported Canadian individual with ALS, who was not known to be related to the USALS#5 family.

**Figure S2.**
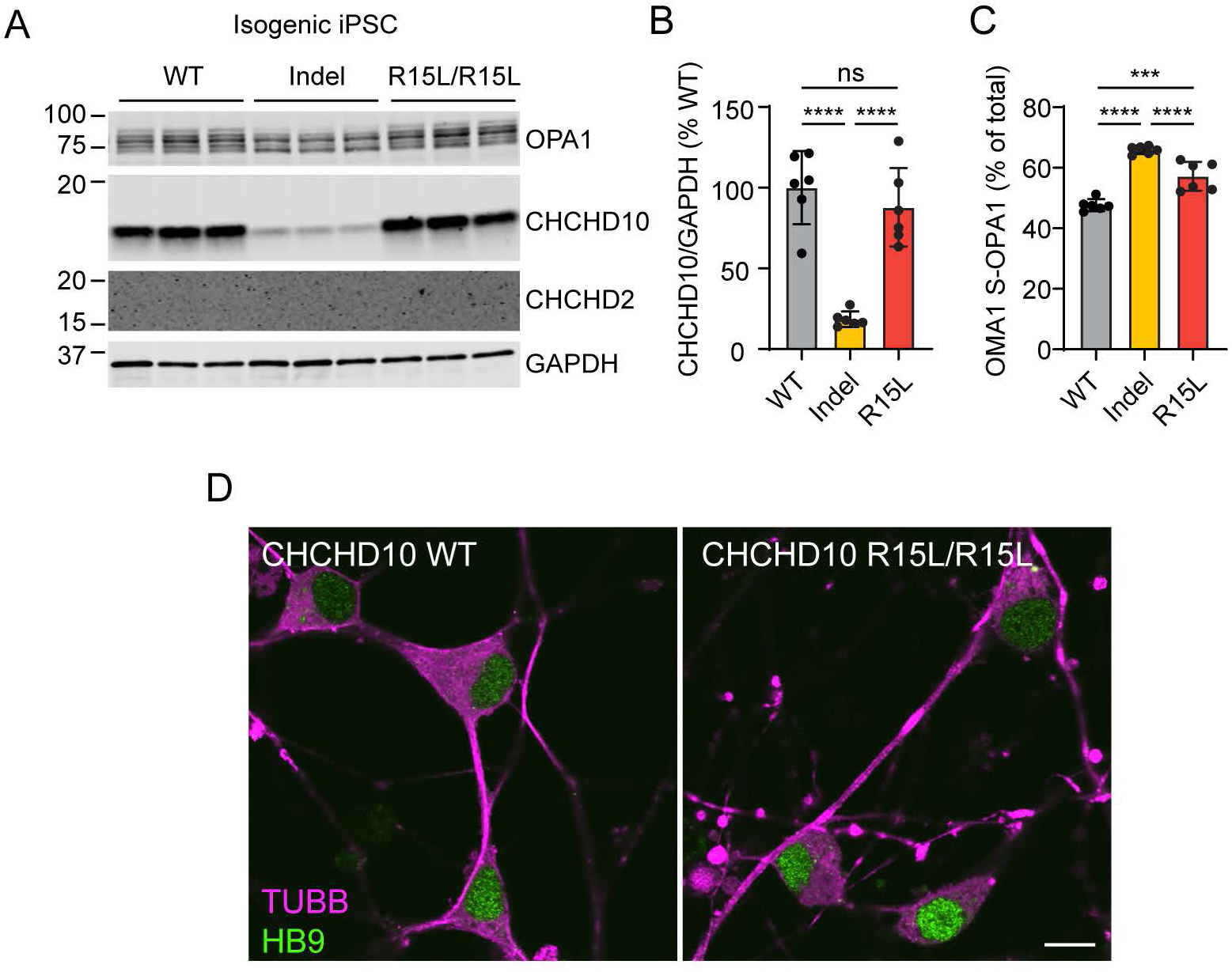
Comparison of indel and R15L/R15L isogenic iPSC cell lines. (A and B) CHCHD10 protein levels (A and B) and OPA1 cleavage by OMA1 (A and C) were measured by immunoblotting for isogenic WT, *CHCHD10* indel, and *CHCHD10* R15/R15L iPSC cell lines. OMA1 activation is reflected by the cleavage of L-OPA1 (*a* and *b* isoforms) to specific S-OPA1 isoforms (*c* and *e* isoforms). (D) iLMNs differentiated as in (Fig. 3H) were immunostained for lower motor neuron markers TUBB (magenta) and HB9 (green). Wells from the same differentiation as (Fig. 3H) were used. In all panels, ns, *, **, ***, ***” correspond to not-significant, p ≤ 0.05, p ≤ 0.01, p ≤ 0.001, and p ≤ 0.0001, respectively.

**Figure S3.**
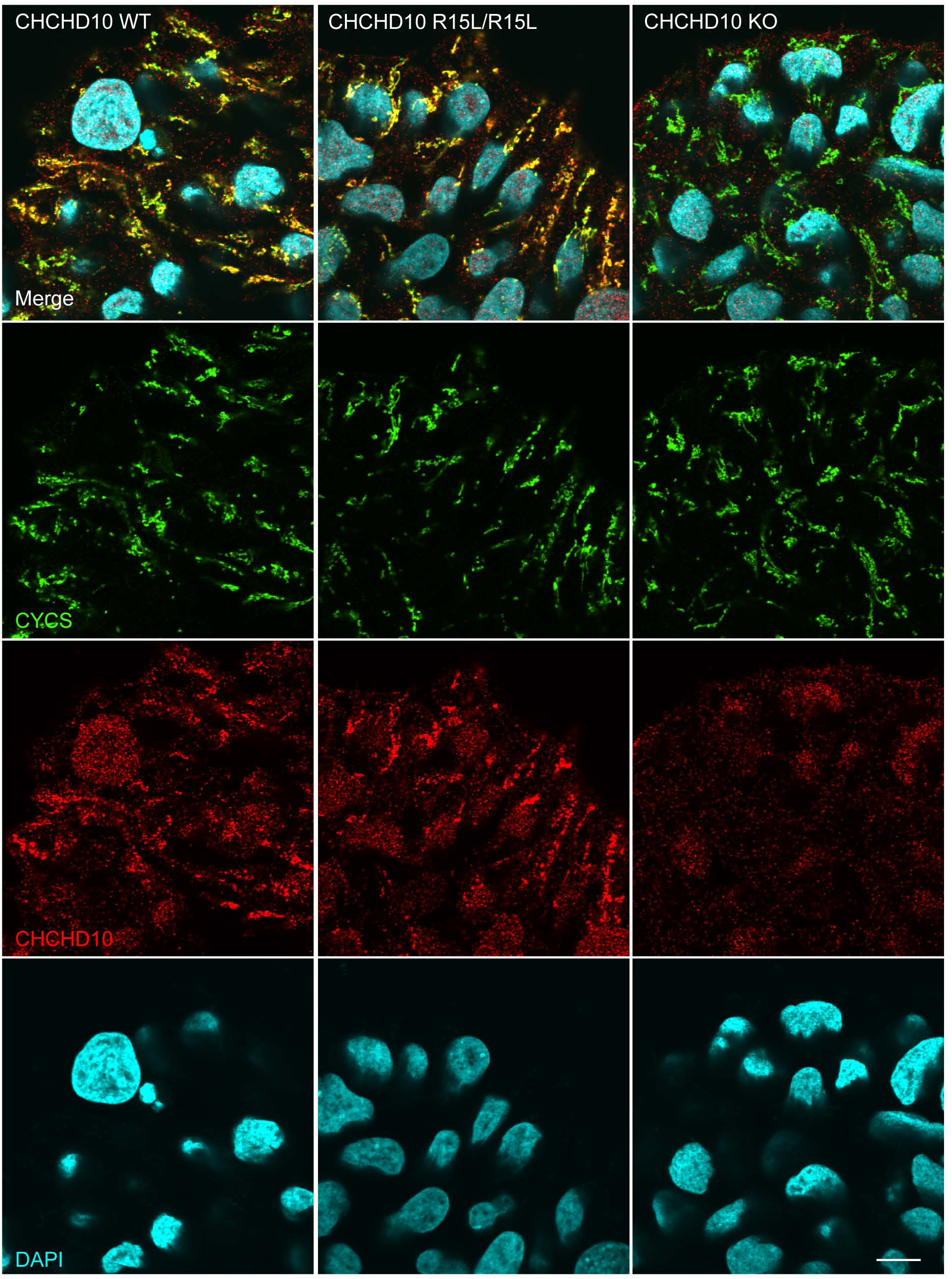
CHCHD10 R15L subcellular localization in iPSC cells. The subcellular localization of CHCHD10 (red) in isogenic parent, R15L/R15L, KO iPSC cells was determined by immunocytochemistry. CHCHD10 co-localized with the mitochondrial marker cytochrome *c* (CYCS) in both parent and R15L/R15L cells. Some CHCHD10 background staining was present in the nucleus and cytosol in all three lines including the KO. Scale bar = 10 μm.

**Figure S4.**
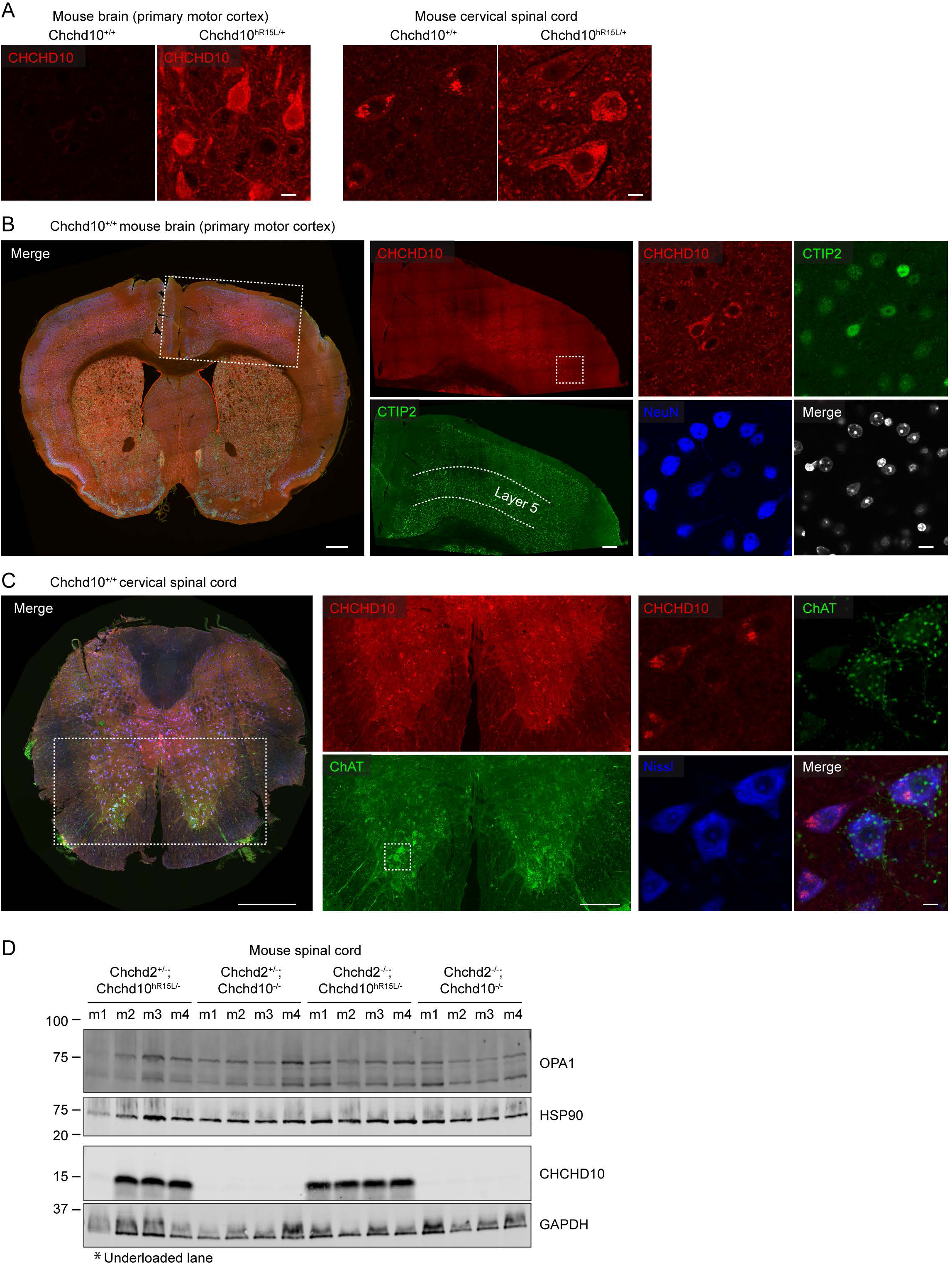
CHCHD10 R15L expression in mouse brain and spinal cord. (A) Comparison of CHCHD10 immunostaining intensity (red) between *Chchd10^hR15L/+^* and *Chchd10^+/+^*brains and spinal cords. Images were windowed the same and the same acquisition parameters were used. The more intense staining in *Chchd10^hR15L/+^*is likely due to higher affinity of the CHCHD10 antibody for human CHCHD10. The images shown here also appear in (Fig. 4C - D and Fig. S4B – C). They are duplicated here to directly compare the CHCHD10 intensity between human CHCHD10 R15L and WT mouse CHCHD10. (B) Distribution of CHCHD10 is shown in the brain of a *Chchd10^+/+^* mouse. WT mouse CHCHD10 has a similar distribution as observed for *Chchd10^hR15L/+^* in (Fig. 4C), although the signal is less intense. (C) Distribution of CHCHD10 is shown in the cervical spinal cord of a *Chchd10^+/+^*mouse. Magnification in right most panels is a z-projection of an image stack. WT mouse CHCHD10 has a similar distribution as observed for *Chchd10^hR15L/+^* in (Fig. 4D), although the signal is less intense. (D) Immunoblotting of CHCHD10 from whole spinal cord lysates from the indicated genotypes. * indicates an underloaded lane, which is apparent also in lower levels of the loading controls HSP90 and GAPDH.

**Figure S5.**
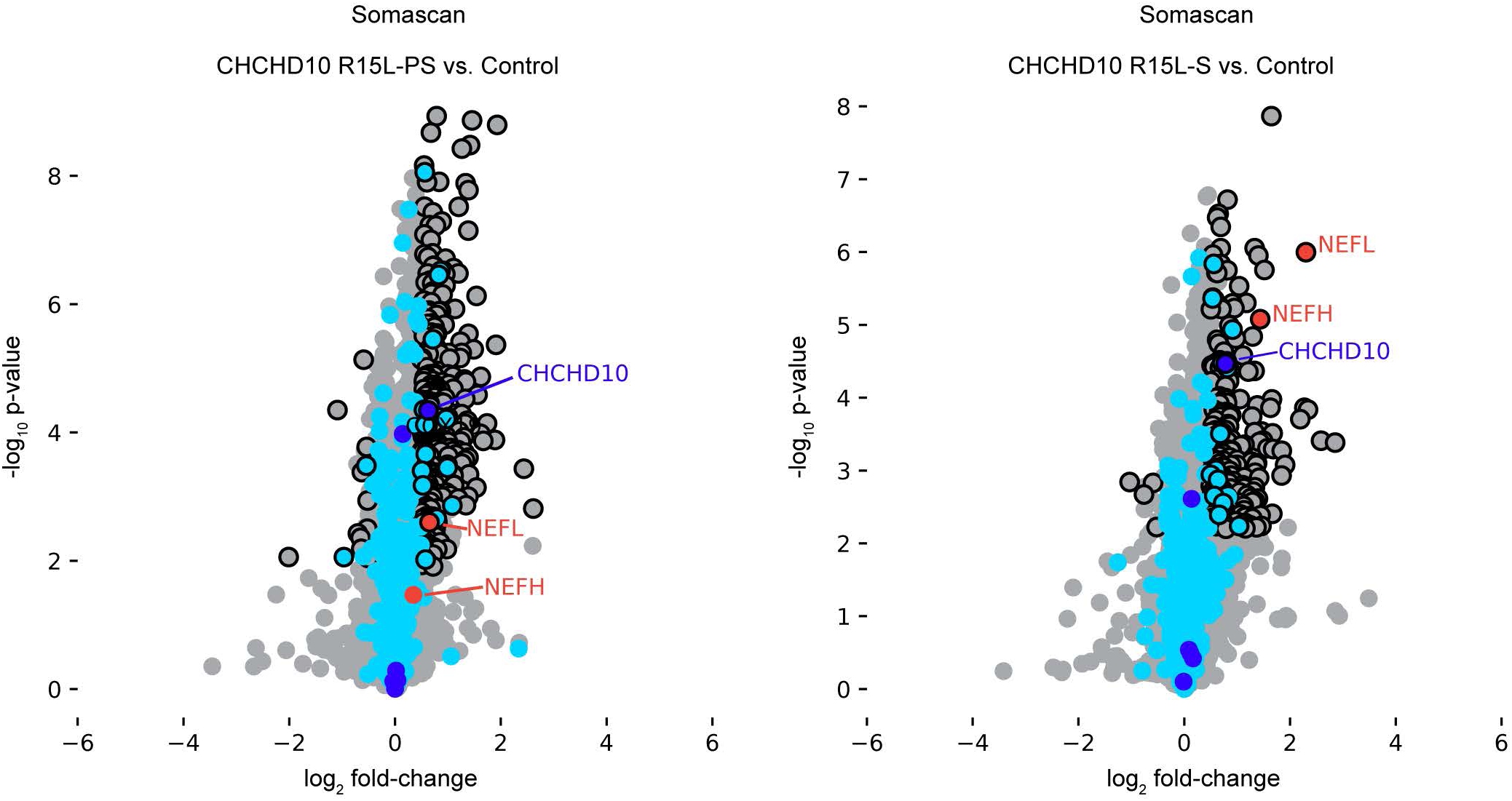
CSF proteomics using the Somascan platform. Volcano plots of CSF proteomics with the Somascan platform comparing symptomatic CHCHD10 R15L patients (C10 R15L – S) to healthy controls (left) and to presymptomatic (C10 R15L – PS) mutation carriers to healthy controls (right). Uncorrected p-value is shown on the y-axis. Significant differentially expressed proteins (FDR < 5% in the one-way ANOVA across all groups and the t-test for the comparison of interest) are outlined in black. Mitochondrial proteins are blue with CHCH-domain containing proteins shown in dark blue and others in light blue.

## Supplemental Tables

**Table S1. Analysis of sibling groups in pedigree.**

**Table S2. Affinity Purification Mass Spectrometry Proteomics.**

**Table S3. Analysis of loss of function CHCHD10 variants in UK Biobank.**

**Table S4. Olink proteomics of CSF.**

**Table S5. Somascan proteomics of CSF.**

## References

1. Feldman EL, Goutman SA, Petri S, et al. Amyotrophic lateral sclerosis. Lancet. 2022;400(10360):1363-1380. doi:10.1016/S0140-6736(22)01272-7

2. Rowland LP, Shneider NA. Amyotrophic lateral sclerosis. N Engl J Med. 2001;344(22):1688-1700. doi:10.1056/NEJM200105313442207

3. Miller TM, Cudkowicz ME, Genge A, et al. Trial of Antisense Oligonucleotide Tofersen for SOD1 ALS. N Engl J Med. 2022;387(12):1099–1110. doi:10.1056/NEJMoa2204705

4. Korobeynikov VA, Lyashchenko AK, Blanco-Redondo B, Jafar-Nejad P, Shneider NA. Antisense oligonucleotide silencing of FUS expression as a therapeutic approach in amyotrophic lateral sclerosis. Nat Med. 2022;28(1):104–116. doi:10.1038/s41591-021-01615-z

5. Shneider NA, Harms MB, Korobeynikov VA, et al. Antisense oligonucleotide jacifusen for FUS-ALS: an investigator-initiated, multicentre, open-label case series. Lancet. 2025;405(10494):2075–2086. doi:10.1016/S0140-6736(25)00513-6

6. Akçimen F, Lopez ER, Landers JE, et al. Amyotrophic lateral sclerosis: translating genetic discoveries into therapies. Nat Rev Genet. 2023;24(9):642–658. doi:10.1038/s41576-023-00592-y

7. Bannwarth S, Ait-El-Mkadem S, Chaussenot A, et al. A mitochondrial origin for frontotemporal dementia and amyotrophic lateral sclerosis through CHCHD10 involvement. Brain. 2014;137(Pt 8):2329–2345. doi:10.1093/brain/awu138

8. Müller K, Andersen PM, Hübers A, et al. Two novel mutations in conserved codons indicate that CHCHD10 is a gene associated with motor neuron disease. Brain. 2014;137(Pt 12):e309. doi:10.1093/brain/awu227

9. Johnson JO, Glynn SM, Gibbs JR, et al. Mutations in the CHCHD10 gene are a common cause of familial amyotrophic lateral sclerosis. Brain. 2014;137(Pt 12):e311. doi:10.1093/brain/awu265

10. Kurzwelly D, Krüger S, Biskup S, Heneka MT. A distinct clinical phenotype in a German kindred with motor neuron disease carrying a CHCHD10 mutation. Brain. 2015;138(Pt 9):e376. doi:10.1093/brain/awv014

11. Penttilä S, Jokela M, Bouquin H, Saukkonen AM, Toivanen J, Udd B. Late onset spinal motor neuronopathy is caused by mutation in CHCHD10. Ann Neurol. 2015;77(1):163–172. doi:10.1002/ana.24319

12. Ajroud-Driss S, Fecto F, Ajroud K, et al. Mutation in the novel nuclear-encoded mitochondrial protein CHCHD10 in a family with autosomal dominant mitochondrial myopathy. Neurogenetics. 2015;16(1):1–9. doi:10.1007/s10048-014-0421-1

13. Shammas MK, Huang X, Wu BP, et al. OMA1 mediates local and global stress responses against protein misfolding in CHCHD10 mitochondrial myopathy. J Clin Invest. Published online June 14, 2022:e157504. doi:10.1172/JCI157504

14. Anderson CJ, Bredvik K, Burstein SR, et al. ALS/FTD mutant CHCHD10 mice reveal a tissue-specific toxic gain-of-function and mitochondrial stress response. Acta Neuropathol. 2019;138(1):103–121. doi:10.1007/s00401-019-01989-y

15. Harjuhaahto S, Jokela M, Rajendran J, et al. Dose-dependent CHCHD10 dysregulation dictates motor neuron disease severity and alters creatine metabolism. Acta Neuropathol Commun. 2025;13(1):111. doi:10.1186/s40478-025-02039-3

16. Keith JL, Swinkin E, Gao A, et al. Neuropathologic description of CHCHD10 mutated amyotrophic lateral sclerosis. Neurol Genet. 2020;6(1):e394. doi:10.1212/NXG.0000000000000394

17. Shammas MK, Huang TH, Narendra DP. CHCHD2 and CHCHD10-related neurodegeneration: molecular pathogenesis and the path to precision therapy. Biochem Soc Trans. 2023;51(2):797–809. doi:10.1042/BST20221365

18. Burstein SR, Valsecchi F, Kawamata H, et al. In vitro and in vivo studies of the ALS-FTLD protein CHCHD10 reveal novel mitochondrial topology and protein interactions. Hum Mol Genet. 2018;27(1):160–177. doi:10.1093/hmg/ddx397

19. Nguyen MK, McAvoy K, Liao SC, et al. Mouse midbrain dopaminergic neurons survive loss of the PD-associated mitochondrial protein CHCHD2. Hum Mol Genet. 2022;31(9):1500–1518. doi:10.1093/hmg/ddab329

20. Liu YT, Huang X, Nguyen D, et al. Loss of CHCHD2 and CHCHD10 activates OMA1 peptidase to disrupt mitochondrial cristae phenocopying patient mutations. Hum Mol Genet. Published online April 27, 2020. doi:10.1093/hmg/ddaa077

21. Genin EC, Madji Hounoum B, Bannwarth S, et al. Mitochondrial defect in muscle precedes neuromuscular junction degeneration and motor neuron death in CHCHD10S59L/+ mouse. Acta Neuropathol. 2019;138(1):123–145. doi:10.1007/s00401-019-01988-z

22. Woo JAA, Liu T, Trotter C, et al. Loss of function CHCHD10 mutations in cytoplasmic TDP-43 accumulation and synaptic integrity. Nat Commun. 2017;8:15558. doi:10.1038/ncomms15558

23. Straub IR, Janer A, Weraarpachai W, et al. Loss of CHCHD10-CHCHD2 complexes required for respiration underlies the pathogenicity of a CHCHD10 mutation in ALS. Hum Mol Genet. 2018;27(1):178–189. doi:10.1093/hmg/ddx393

24. Brockmann SJ, Freischmidt A, Oeckl P, et al. CHCHD10 mutations p.R15L and p.G66V cause motoneuron disease by haploinsufficiency. Hum Mol Genet. 2018;27(4):706–715. doi:10.1093/hmg/ddx436

25. Ryan ÉB, Yan J, Miller N, et al. Early death of ALS-linked CHCHD10-R15L transgenic mice with central nervous system, skeletal muscle, and cardiac pathology. iScience. 2021;24(2):102061. doi:10.1016/j.isci.2021.102061

26. Liu T, Woo JAA, Bukhari MZ, et al. Modulation of synaptic plasticity, motor unit physiology, and TDP-43 pathology by CHCHD10. Acta Neuropathol Commun. 2022;10(1):95. doi:10.1186/s40478-022-01386-9

27. van Rheenen W, Diekstra FP, van den Berg LH, Veldink JH. Are CHCHD10 mutations indeed associated with familial amyotrophic lateral sclerosis? Brain. 2014;137(Pt 12):e313. doi:10.1093/brain/awu299

28. Staffaroni AM, Asken BM, Casaletto KB, et al. Development and validation of the Uniform Data Set (v3.0) executive function composite score (UDS3-EF). Alzheimers Dement. 2021;17(4):574–583. doi:10.1002/alz.12214

29. Samra K, MacDougall AM, Peakman G, et al. Motor symptoms in genetic frontotemporal dementia: developing a new module for clinical rating scales. J Neurol. 2023;270(3):1466–1477. doi:10.1007/s00415-022-11442-y

30. Snyder et al. Alzheimer and Dementia. Published online 2025.

31. Cedarbaum JM, Stambler N, Malta E, et al. The ALSFRS-R: a revised ALS functional rating scale that incorporates assessments of respiratory function. BDNF ALS Study Group (Phase III). J Neurol Sci. 1999;169(1-2):13–21. doi:10.1016/s0022-510x(99)00210-5

32. Fischl B, Sereno MI, Dale AM. Cortical surface-based analysis. II: Inflation, flattening, and a surface-based coordinate system. Neuroimage. 1999;9(2):195–207. doi:10.1006/nimg.1998.0396

33. Fischl B, Salat DH, Busa E, et al. Whole brain segmentation: automated labeling of neuroanatomical structures in the human brain. Neuron. 2002;33(3):341–355. doi:10.1016/s0896-6273(02)00569-x

34. Fischl B, van der Kouwe A, Destrieux C, et al. Automatically parcellating the human cerebral cortex. Cereb Cortex. 2004;14(1):11–22. doi:10.1093/cercor/bhg087

35. Desikan RS, Ségonne F, Fischl B, et al. An automated labeling system for subdividing the human cerebral cortex on MRI scans into gyral based regions of interest. Neuroimage. 2006;31(3):968–980. doi:10.1016/j.neuroimage.2006.01.021

36. Malone IB, Leung KK, Clegg S, et al. Accurate automatic estimation of total intracranial volume: a nuisance variable with less nuisance. Neuroimage. 2015;104:366–372. doi:10.1016/j.neuroimage.2014.09.034

37. Karczewski KJ, Francioli LC, Tiao G, et al. The mutational constraint spectrum quantified from variation in 141,456 humans. Nature. 2020;581(7809):434–443. doi:10.1038/s41586-020-2308-7

38. Chia R, Moaddel R, Kwan JY, et al. A plasma proteomics-based candidate biomarker panel predictive of amyotrophic lateral sclerosis. Nat Med. Published online August 19, 2025. doi:10.1038/s41591-025-03890-6

39. Rath S, Sharma R, Gupta R, et al. MitoCarta3.0: an updated mitochondrial proteome now with sub-organelle localization and pathway annotations. Nucleic Acids Res. 2021;49(D1):D1541–D1547. doi:10.1093/nar/gkaa1011

40. Hu MT, Ellis CM, Al-Chalabi A, Leigh PN, Shaw CE. Flail arm syndrome: a distinctive variant of amyotrophic lateral sclerosis. J Neurol Neurosurg Psychiatry. 1998;65(6):950–951. doi:10.1136/jnnp.65.6.950

41. Kwan JY, Jeong SY, Van Gelderen P, et al. Iron accumulation in deep cortical layers accounts for MRI signal abnormalities in ALS: correlating 7 tesla MRI and pathology. PLoS One. 2012;7(4):e35241. doi:10.1371/journal.pone.0035241

42. Wendebourg MJ, Kesenheimer E, Sander L, et al. The Lateral Corticospinal Tract Sign: An MRI Marker for Amyotrophic Lateral Sclerosis. Radiology. 2024;312(3):e231630. doi:10.1148/radiol.231630

43. Lin HP, Petersen JD, Gilsrud AJ, et al. DELE1 maintains muscle proteostasis to promote growth and survival in mitochondrial myopathy. EMBO J. 2024;43(22):5548–5585. doi:10.1038/s44318-024-00242-x

44. Markouli C, Couvreu De Deckersberg E, Regin M, et al. Gain of 20q11.21 in Human Pluripotent Stem Cells Impairs TGF-β-Dependent Neuroectodermal Commitment. Stem Cell Reports. 2019;13(1):163–176. doi:10.1016/j.stemcr.2019.05.005

45. Zhu L, Gomez-Duran A, Saretzki G, et al. The mitochondrial protein CHCHD2 primes the differentiation potential of human induced pluripotent stem cells to neuroectodermal lineages. J Cell Biol. 2016;215(2):187–202. doi:10.1083/jcb.201601061

46. Kim J, Kwon EJ, Kim YJ, et al. Epigenetic repression of CHCHD2 enhances survival from single cell dissociation through attenuated Rho A kinase activity. Cell Mol Life Sci. 2024;81(1):38. doi:10.1007/s00018-023-05060-8

47. Gao J, Douglas AGL, Chalitsios CV, et al. Neurodegenerative disease in C9orf72 repeat expansion carriers: population risk and effect of UNC13A. Brain. Published online July 19, 2025:awaf269. doi:10.1093/brain/awaf269

48. Petzold A. The 2022 Lady Estelle Wolfson lectureship on neurofilaments. J Neurochem. 2022;163(3):179–219. doi:10.1111/jnc.15682

49. Oeckl P, Jardel C, Salachas F, et al. Multicenter validation of CSF neurofilaments as diagnostic biomarkers for ALS. Amyotroph Lateral Scler Frontotemporal Degener. 2016;17(5-6):404–413. doi:10.3109/21678421.2016.1167913

50. Han X, Zhan F, Yao Y, Cao L, Liu J, Yao S. Clinical heterogeneity in a family with flail arm syndrome and review of hnRNPA1-related spectrum. Ann Clin Transl Neurol. 2022;9(12):1910–1917. doi:10.1002/acn3.51682

51. Liu Q, Shu S, Wang RR, et al. Whole-exome sequencing identifies a missense mutation in hnRNPA1 in a family with flail arm ALS. Neurology. 2016;87(17):1763–1769. doi:10.1212/WNL.0000000000003256

52. Riva N, Pozzi L, Russo T, et al. NEK1 Variants in a Cohort of Italian Patients With Amyotrophic Lateral Sclerosis. Front Neurosci. 2022;16:833051. doi:10.3389/fnins.2022.833051

53. Kirby J, Goodall EF, Smith W, et al. Broad clinical phenotypes associated with TAR-DNA binding protein (TARDBP) mutations in amyotrophic lateral sclerosis. Neurogenetics. 2010;11(2):217–225. doi:10.1007/s10048-009-0218-9

54. Solski JA, Yang S, Nicholson GA, et al. A novel TARDBP insertion/deletion mutation in the flail arm variant of amyotrophic lateral sclerosis. Amyotroph Lateral Scler. 2012;13(5):465–470. doi:10.3109/17482968.2012.662690

55. Phukan J, Pender NP, Hardiman O. Cognitive impairment in amyotrophic lateral sclerosis. Lancet Neurol. 2007;6(11):994–1003. doi:10.1016/S1474-4422(07)70265-X

56. Lv G, Sayles NM, Huang Y, et al. Amyloid fibril structures link CHCHD10 and CHCHD2 to neurodegeneration. Nat Commun. 2025;16(1):7121. doi:10.1038/s41467-025-62149-3

57. Fisher EMC, Greensmith L, Malaspina A, et al. Opinion: more mouse models and more translation needed for ALS. Mol Neurodegener. 2023;18(1):30. doi:10.1186/s13024-023-00619-2

58. Wittenbach JD, Cocanougher BT, Yun P, Foley AR, Bönnemann CG. MuscleViz: Free Open-Source Software for Muscle Weakness Visualization. J Neuromuscul Dis. 2019;6(2):263–266. doi:10.3233/JND-190385

59. Woolley SC, York MK, Moore DH, et al. Detecting frontotemporal dysfunction in ALS: utility of the ALS Cognitive Behavioral Screen (ALS-CBS). Amyotroph Lateral Scler. 2010;11(3):303–311. doi:10.3109/17482961003727954

