## Supplemental Text for "Clinical, neuropathological, and biochemical characterization of ALS in a large CHCHD10 R15L family"

**Guide RNAS and repair ssODN donor sequences used to generate isogenic KO and KI iPSC lines:**

| Isogenic Variant | sgRNA | ssODN |
| --- | --- | --- |
| *CHCHD10* c.270-299del; c.288delA | CCCGCUGAAGGCUCCGGUCA | GCAGGGGTAGCCGTGGGCTCGGCTGTGGGACACGTCATGGTCTCCGCTCTGACCGGAGCCTTCAGCGGGGGGAGCTCGGAGCCCTCCC |
| *CHCHD10* p.R15L | AGGGCGCGGCGGGGCGGCUG | CCAGCCTGGGGTCTGGCCAGGATTAACCCTGCTTCCTCCCACCCCCGCAGCCTCCCCGCCGCGCCCTCTGCCCACCCGCCCGCGCACCCACCGCCCTCGG |
| *CHCHD10* KO | UCGGCUGUGGGACACGUCAU |  |

**Sequencing primers used for Sanger sequencing of CHCHD10 clones for haplotype analysis:**

| **Primer Name** | **Primer Sequence** |
| --- | --- |
| Primer 1 | AGAGATGGACGACCCACGTC |
| Primer 2 | CTTTGTGGCTAGGTCAGGGG |
| Primer 3 | TGGAGGTGCGTTTCAAACCT |
| Primer 4 | GGGGCTCAGGAAATCCAACATC |
| Primer 5 | TCGAGGCCACACAGGGAG |
| Primer 6 | AATTTGTTGGGCATGGTGGC |
| Primer 7 | AGACCAGTGGACTAGGACCC |
| Primer 8 | TCAAGTGGTTGTCCTGCC |
| Primer 9 | TCGAGAGAGGCACACAACAG |
| Primer 10 | GGCTGAAGCAATGGTGAATAC |
| Standard Primer M13R | CAGGAAACAGCTATGACC |
| Standard Primer T7 | TAATACGACTCACTATAGGG |
| Standard Primer T3 | AATTAACCCTCACTAAAGGG |

**Amplification primers used for long range PCR of CHCHD10 for cloning of each CHCHD10 allele:**

| **Primer Name** | **Primer Sequence** |
| --- | --- |
| Forward Primer | CTCTCCACCTCACTCTGGACTG |
| Reverse Primer | GAGCAGGGTCCCATTTCCAG |

**Donor vector sequence for Chchd10^R15L/+^ mouse line:**

GGATCCagccagaccccccaccccccccaccctccacttcccacccccttaccccccaacctgccccattaccccacattaccacacttagtagaggatctggaaggtgggctagccacggaggcccacagaaggggttgaagctggaagaatgttaccacttgcctactggatacaaggtaagcagaggccaaatagataagccagtggggtctgctcaagcctgcccccccctttttttttttaattttctacaacagcacatttttttaaaagatttatttatttattatatgtaagtacactgtagctgtcttcagacactcaagaagagggcatcagatttcgttatggatggttgtgagccaccatgtggttgctgggatttgaactcgggaccttcagaagagcagtcggcactcttaaccactgagccatctcgccagccccctgcccccccttttttaaaagatgtatttatttatttcgtgtatgtgggtacattgtcgctgccttcagacacactagaagagggcatcagatctcattatagacagttgtgagccaccatgtagttgctgggaattgaactcaggacctctggaagagcagacagctctcataaccactgaaccatctctctccagccccagtcagtgctcttaacagttaagccacctctctagcctgagtctggcatttttttaaaggaaagagtggcttgtcttttacctactcacctcaccaaaaagtctatcctgcagatgggtggtgattccacttgggaggaaagctgaagtcttaatgaaagaggcctgctacagaggctgcttaaagaaaccaggtttgggacacctgaatgggcaatgcatggtcctgaatagttttgacccctgttgctgaggcatttgaattgtttgtgccaagtgatctctgttgtcacagtaaaataaacactgaaagaagaggagaaagaactcaaggccccctgtgccctttctactctggccaaaggttattattatttgttttattttgttttgttttgtgggagacagttctgtgtagccctggaactcagtctgtagaccaggctggcctcgaacacacaagatgcacctacctatacctgccccccccccaagttctggggttacaggtgtgtgctcttttaaccttttttttttttttctctctccagAGGTTATGGCATGATCCCAGCCAAGGTCTCTGAGGAAAAGTCTGTGAAAACCGAGTTCCAGAGGACAGGGAAGGGCCTTTGGTGATCCTGGGATGGTTATTTTCAAACTCTGTCTTGTGATGGGTCTCAGAGAACACAACCTGGCCTGCAGTCTCCATTCGCTTCTCCGCTGGCGGTCTATGTGTCTCTTTCTTTCCTCCTGGGCTCGTGGCAAACCCCTATTGCATTGCTTGAGTTCTGACAGAAAGACCTGCCCTCGCTGCCCCGTGCCTAACTAAGCCTTGGGCTTTCCAAAGCTGACCGGTGTCCCCAGGACTCGCGTAGCTCCGCCCTGCACTGGGATAGCTCCGGAGCCCTAAATCCAGGAGGTTAATGGAAGGGCGAGCTTTGGCCCCTCCCATTTGGAGGGCGTGGATCAGAGGCCCCGCCCCAGATCCCGCCCATTTCACCTGACTCTAAGGACGCTGCCGTGTTAGTTGCCCGGCACTGAGCGAGCGGCTGCTACGGTCCACGCTTCTTACTCAGCTCCAACTTGATCCCACCGCAGTCATGCCTCGGGGAAGCCGCAGCGCGGCCTCCCGGCCAGCCAGgtgtgagggggacaagggccctgtggggagggggttagggaagtgggggcactcccctaagggcgtccgcagagtgaccttggctgctgctgcgccatcttggcggggggaggggtgggggataacttcgtataatgtatgctatacgaagttattgcagcacccacgtgtggagacgtgggtcgtccatctctccgggggtcgtttccaggagctgcagccctccagcctggggtctggccaggattaaccctgcttcctcccacccccgcagCCTCCCCGCCGCGCCCTCTGCCCACCCGCCCGCGCACCCACCGCCCTCGGCAGCCGCCCCAGCCCCCGCCCCTTCGGGCCAGCCGGGGCTCATGGCTCAGATGGCGACCACGGCCGCAGGGGTAGCCGTGGGCTCGGCTGTGGGACACGTCATGGGCAGCGCCCTGACCGGAGCCTTCAGCGGGGGGAGCTCGGAGCCCTCCCAGCCTGCTGTCCAGCAGgtgagcagcgggtccaagagaaactgaggcaggattatctcgaggccacacagggagctgcccaagtgtccagtgaggagtgtccgcgtgtattcaccattgcttcagcccaggactctgcccgggaagggtcctagtccactggtcttgagttcactgtcacttagaggcaggcttcccggcctctcccttgaggccagggtccctaagctcccagggagagacaccaaccagccctttaaagatgggaaacccaggccctgagggctcagagactcgcccaaggtcacctcaccctagcagaggggagggatgttggatttcctgagcccctgttctccccacacccaggtctgggcactgtggccgctgggcgcaagggcgaaggggagccataacttcgtataatgtatgctatacgaagttatacttccctgatgacctttcccctgctaggaaccccaccgccgtaagccaaatgtggctttgtggctaggtcaggggtcatgaggattagtgtagctgagccaggaaggtgctagaacttgtgggggtggggctcatactctgagagtacaggcagagtactgggctttcctagagccttctcctccctcccctgcacccgcccaaagtgcccattagaaggcagcagaggccaggcgcagtggctcatgcctgtaatcccagcactttgggaggccaaggcgggtggatcatgagttcaggagttcgataccagcctggccaacatagtgaaaccccctctctactaaaaatacaaaaaatttgttgggcatggtggcgggcgcctgtagccccagctacttgggaggccgaggcaggacaaccacttgaacccgggaggcggaggttgcagtgagccgagactgcgccgctgcactccactctaggtgacagagtgagactctgtctcaaaaaaaaaaaaaaaaaaaggcagcagaaacatgctgtctgtagccgcctctgctgctggatcactcagcctgcctaggacccttgggacacccaggctggggatgtggggaggggcaggtggccccaggtttgaaacgcacctccaggtgccaactccaagctgatcctgcagcctcttgcactgtacccccagGCCCCCACCCCCGCTGCCCCCCAGCCCCTGCAGATGGGGCCCTGCGCCTATGAGATCAGGCAGTTCCTGGACTGTTCCACCACTCAGAGTGACCTGTCCCTGTGTGAGGGCTTCAGCGAGGCCCTGAAGCAGTGCAAGTACTACCATGgtgagtgagtggaccccgactcaggccgggagggggagaggccaaccctcctcttgcacctgcaggctgaccaccagcctgtgctgcccctcccttcccagGTCTGAGCTCCCTGCCCTGAAGAGGCTGTGGGCCTGCTCATCGCCTAACCCCTCACCGACAGCTTGATGGAAAGGAGAGGTTCCATGTGACTGGGAGTAAGGAAGGATCGTTCTCACCCCGCAGACTAACAGGAGACATAATTATTCAATTAAAAAGTTTACTTTAGACCACAGCCtgctgtgtgcccatttactgccctgttggctggcgggggggggggggggggggcaggatcaggggagataggggaacagctatcttgttaatgtgctatgcctcatctttgcggaggtgggagagaggatgtgggagttggggatcttgttatagaaccctgagctgtccttgaacctctgggatgggaggaactcctatctgtctgcattgtggatagtcaaggacactcctaagcagggagactggagaggtatttgcaagtggtcccctgggacctaaaacctcaagaggagcctattgtctctgcccacaggaacatctgtgtcttttctcttcttattttgtccttgagttcttttttctgcactcaaatggggtggggtggggatctcaagcgctcggggagtggggggggggcaactctcacatcctgtctgtgacctttcccttcttgctgatgtaacttcttagtaaccgccagatcctggaagaggagtcctgctggattgctgtctgtttgcagttgatcataaagcagaggtcagggccagtccccttggattcaagtccagcttggctgtggtgtaccggctcatagaacctgctggaatggaagcaggcccttaagaccacagctcagaaaaaaaaaaaaaaaagacacagctcagattgcttcatcgtacagtcgctacgttcactgagttcagtcaacaacatgcattgaacacctactgtgtgctaggctggagaaacgtgggccagcttcctaaacgcagagtatgtgtgcctttaggatgctgatagtccagcaatactgtggggttttatctctggttacagtcctgggtgtgtctctgtgtgaaagtcagtaaagatcccttttggattttcctgggttcccctacctttttctttttctctttctttctttctttctttctttctttctttctttctttctttctttctttctttctttctttttttttttttggtttttcgagacagggtttctctgtgtagccctgctgtcctggaactcactttgtagaccaggctggcctcgaactcagaaattcgtctgcctctgtctcccaagtgctgggattaaaggcatgtaccaccaccgcctggcaaaatctttatttatctatctatctatctatctatctatctatctatctatctatctatctatctatctatttattaatgtatgcatgtatttagagataggaccaactaagttgctcaggctatctgtctgtctgtctgtctgtctatctatttattaatgtatgtatgtatttagagataggaccaactaagttgctcatgctgtccttgaacttactctatagtccGGATCC
